# The Metabolomic Profile of a Healthy Lifestyle Mediates Psoriasis Risk and Predicts Multiple Comorbidities

**DOI:** 10.1101/2025.07.22.25331968

**Authors:** Shiyu Zhang, Yu Meng, Yuming Sun, Yao Yu, Zehao Luo, Daishi Li, Ziyu Guo, Jinchen Li, Furong Zeng, Guangtong Deng, Xiang Chen

**Author notes:** Correspondence: Guangtong Deng,; Xiang Chen,. Department of Dermatology, Xiangya Hospital, Central South University, Changsha, Hunan, China. Equal contribution (Shiyu Zhang, Yu Meng).

## Abstract

Psoriasis is increasingly recognized as a systemic metabolic condition with complex immunometabolic mechanisms. While a healthy lifestyle is associated with reduced psoriasis risk, it remains unclear whether and how metabolomic changes mediate this association. We leveraged metabolomic data on 327 NMR biomarkers from 275,326 UK Biobank participants to investigate how lifestyle and metabolism relate to psoriasis and its comorbidities. We constructed a healthy lifestyle score incorporating ten modifiable behaviors and identified a comprehensive metabolomic profile comprising 112 NMR features associated with healthy living. This metabolomic profile mediated 37.5% to 46.1% of the total effect of lifestyle on psoriasis risk. Notably, three biomarkers—GlycA, PUFA/MUFA ratio, and creatinine—were identified as key mediators with shared genetic architectures with psoriasis, particularly involving the MHC region, underscoring the pivotal role of T-cell activation and antigen-presentation in the immunometabolic axis of psoriasis and suggesting a novel “metabolic-renal” pathway contributing to disease progression. Furthermore, maintaining a healthy lifestyle and a favorable metabolomic profile helped prevent severe comorbidities in individuals with psoriasis, with GlycA emerging as a promising prognostic biomarker for future clinical application. These findings establish a mechanistic connection between lifestyle, metabolism, and psoriasis pathogenesis, highlighting metabolism-oriented lifestyle modification as a strategy for psoriasis and related comorbidity management.

## Introduction

Psoriasis, a chronic inflammatory skin disease, affects an estimated 100 million people globally[1], imposing a considerable medical burden on healthcare systems worldwide[2, 3]. Beyond its primary dermatological manifestations, emerging epidemiological evidence reveals an extensive comorbidity profile encompassing psoriatic arthritis, cardiovascular diseases (CVDs), metabolic syndrome, and various malignancies[4, 5], which collectively amplify the disease’s clinical burden and significantly deteriorate the quality of life for affected individuals.

Emerging evidence highlights that metabolic dysregulation plays a pivotal role in psoriasis pathogenesis[6–9]. Multiple metabolic pathways—such as glycolysis, the tricarboxylic acid (TCA) cycle, lipid metabolism, and amino acid metabolism—are known to impact both keratinocytes and immune cells[6], underscoring the promise of metabolism-targeted strategies in psoriasis prevention and treatment.

Lifestyle modification is gaining recognition in psoriasis management due to the preventive benefits of healthy behaviors, including dietary choices, smoking and drinking habits, and regular physical activity[10–12]. These modifiable factors impact systemic metabolic networks and contribute to metabolic health[13, 14], supporting the notion that lifestyle-induced metabolic changes could serve as a bridge linking environmental exposures and disease progression[15–17]. However, the specific metabolomic features and pathways through which lifestyle factors affect psoriasis risk remain unknown.

The UK Biobank has measured nuclear magnetic resonance (NMR)-based plasma metabolomic biomarkers in 275,226 participants, alongside rich phenotypic and genotypic data[18]. Leveraging this resource, our study aims to: (1) establish a holistic metabolomic profile associated with healthy living; (2) elucidate how lifestyle and associated metabolomic signatures relate to psoriasis and multiple comorbidities; and (3) identify key metabolomic biomarkers and potential mechanisms underlying psoriasis pathogenesis (**Figure 1**). By uncovering metabolomic links between lifestyle and psoriasis, our study could inform early risk stratification, guide personalized interventions, and facilitate effective clinical translation.

**Figure 1.**
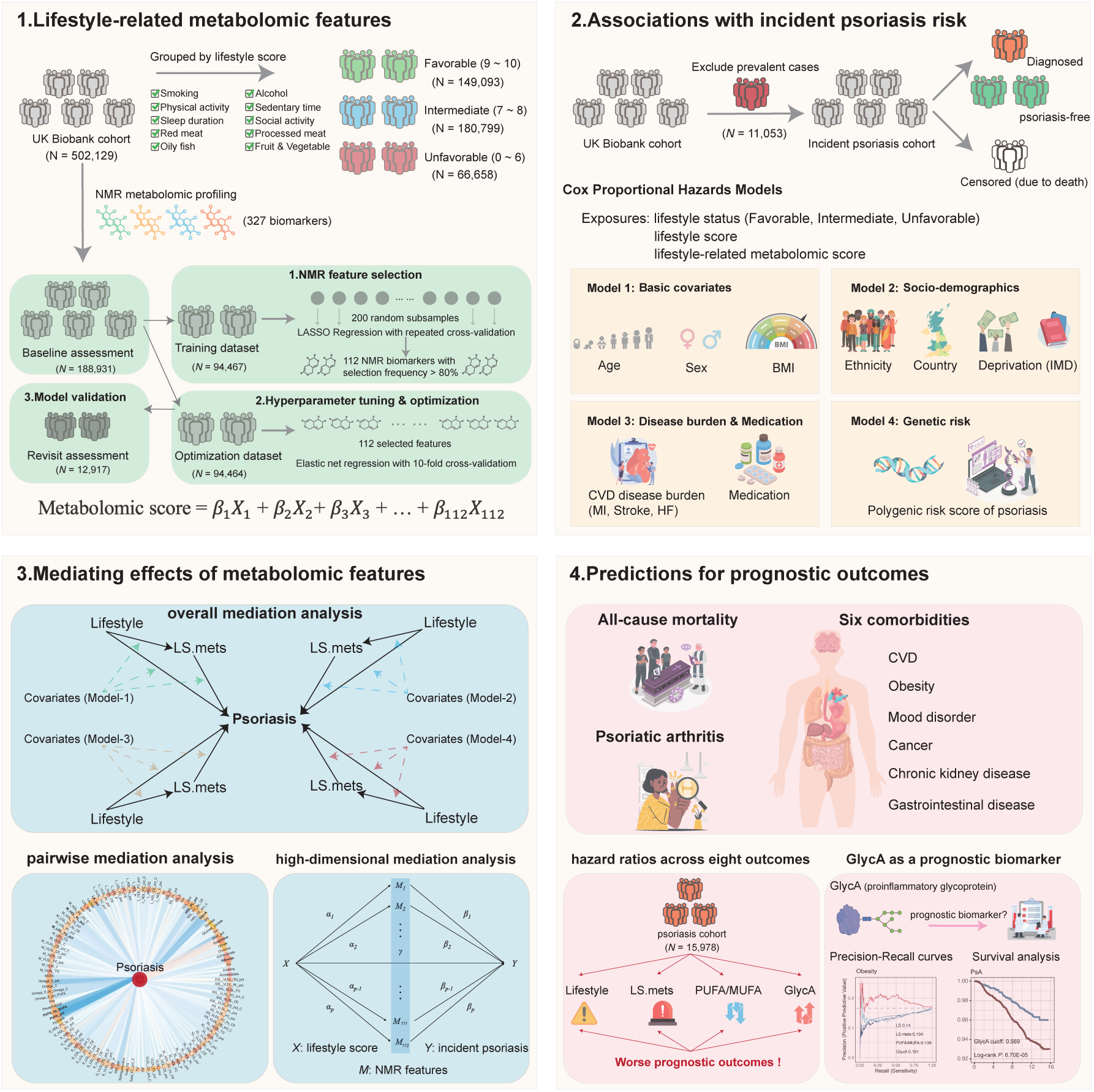
Study overview. (1) A healthy lifestyle score (LS) was constructed from ten lifestyle factors in the UK Biobank. A three-step machine learning approach identified 112 metabolomic features and developed a composite lifestyle-associated metabolomic score (LS.mets). (2) An incident psoriasis cohort (*N*=4,714) was established to investigate the associations of lifestyle and the metabolomic profile with psoriasis risk, using four Cox proportional hazards models adjusted for different sets of confounders. (3) Mediation analyses were conducted to examine the extent to which the LS.mets and individual metabolomic features mediate the relationship between lifestyle and psoriasis. (4) Finally, we evaluated the associations of the LS, LS.mets, and key mediating biomarkers with eight clinical prognostic outcomes in psoriasis patients, highlighting GlycA as a potential novel prognostic biomarker.

## Results

### Identification of a healthy lifestyle-related metabolomic profile

Our study population included 502,129 participants from the UK Biobank cohort, for whom extensive environmental, lifestyle, and genetic data were collected (**Table 1**). A healthy lifestyle pattern was defined based on ten components: smoking status, alcohol consumption, physical activity, sedentary time, sleep duration, fruit and vegetable intake, oily fish intake, red meat intake, processed meat intake, and social activity. A healthy lifestyle score, ranging from 0 to 10, was constructed based on the fulfillment of criteria for these ten components, with a higher score indicating a healthier lifestyle (Methods). Participants were further categorized into three groups based on their scores: unfavorable (0 - 6 points), intermediate (7 - 8 points), and favorable (9 - 10 points). We observed that the distributions of lifestyle scores were highly consistent between the baseline assessment and the first revisit. A large proportion of participants scored between 7 and 9 points, while only a relatively small proportion had an unfavorable lifestyle (**Figure 2a**).

**Figure 2.**
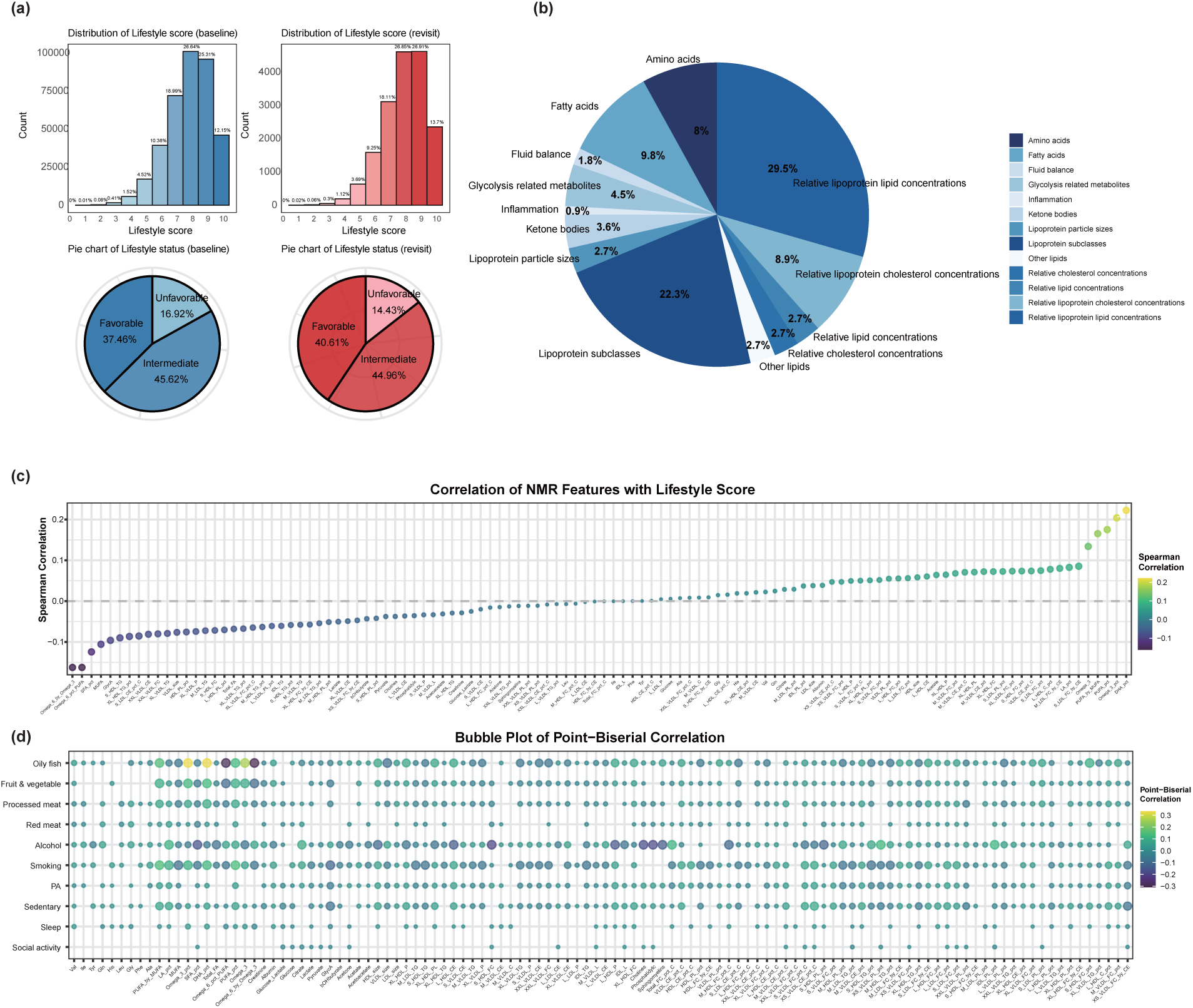
A healthy lifestyle-related metabolomic profile composed of 112 NMR features. (a) Distribution of healthy lifestyle scores at baseline (blue, *N*= 379,376) and at the first revisit (red, *N*= 17,174), alongside pie charts showing the proportions of participants in each lifestyle group: unfavorable (0-6 points), intermediate (7-8 points), and favorable (9-10 points). (b) Pie chart displaying the proportional breakdown of the 112 metabolomic features across 13 categories. (c) Spearman’s correlations of each NMR feature with the lifestyle score (*N*= 201,848), ordered from the smallest to largest coefficient. The color gradient indicates the direction of correlation, and the point size reflects the strength of the correlation, proportional to the negative logarithm of the Bonferroni-corrected p-value. (d) Bubble plot showing the point-biserial correlations of each NMR feature with each lifestyle factor (*N*= 201,848), arranged by their respective categories (see Supplementary Table 1). The color gradient indicates the direction of correlation, and the point size reflects the correlation strength, also proportional to the negative logarithm of the Bonferroni-corrected p-value. Correlations with Bonferroni-corrected p-values > 0.05 are not displayed.

**Table 1.** baseline characteristics of the study population.

| Table1. Baseline characteristics of the study population |  |  |  |  |
| --- | --- | --- | --- | --- |
|  | Sample size (N) | Psoriasis Groups |  |  |
|  |  | Control<br>N = 486,151 | Prevalent<br>N = 11,053 | Incident<br>N = 4,925 |
| Age | 502,129 | 58 (50, 63) | 58 (51, 63) | 59 (51, 64) |
| Sex | 502,129 |  |  |  |
| Male |  | 220,991 (45%) | 5,599 (51%) | 2,383 (48%) |
| Female |  | 265,160 (55%) | 5,454 (49%) | 2,542 (52%) |
| BMI | 499,026 | 26.7 (24.1, 29.9) | 27.5 (24.7, 30.9) | 27.6 (24.9, 31.1) |
| Ethnic background | 499,354 |  |  |  |
| Asian |  | 9,600 (2.0%) | 173 (1.6%) | 99 (2.0%) |
| Black |  | 7,988 (1.7%) | 36 (0.3%) | 24 (0.5%) |
| Chinese |  | 1,548 (0.3%) | 12 (0.1%) | 11 (0.2%) |
| Mixed |  | 2,875 (0.6%) | 54 (0.5%) | 21 (0.4%) |
| Other |  | 4,457 (0.9%) | 62 (0.6%) | 33 (0.7%) |
| White |  | 456,978 (95%) | 10,672 (97%) | 4,711 (96%) |
| Index of multiple deprivation | 489,407 | 13 (7, 23) | 13 (8, 25) | 14 (8, 26) |
| Myocardial infarction | 502,129 |  |  |  |
| Control |  | 474,504 (98%) | 10,680 (97%) | 4,764 (97%) |
| Case |  | 11,647 (2.4%) | 373 (3.4%) | 161 (3.3%) |
| Stroke | 502,129 |  |  |  |
| Control |  | 478,658 (98%) | 10,823 (98%) | 4,824 (98%) |
| Case |  | 7,493 (1.5%) | 230 (2.1%) | 101 (2.1%) |
| Heart failure | 502,129 |  |  |  |
| Control |  | 483,330 (99%) | 10,960 (99%) | 4,880 (99%) |
| Case |  | 2,821 (0.6%) | 93 (0.8%) | 45 (0.9%) |
| Cholesterol lowering medication | 502,129 | 83,465 (17%) | 2,296 (21%) | 1,093 (22%) |
| PRS of psoriasis | 485,904 | -0.35 (-0.86, 0.28) | 0.36 (-0.38, 1.44) | -0.07 (-0.63, 0.72) |
| Smoking status | 379,376 |  |  |  |
| 0 |  | 36,168 (9.8%) | 1,131 (14%) | 548 (15%) |
| 1 |  | 331,319 (90%) | 7,180 (86%) | 3,030 (85%) |
| Alcohol consumption | 379,376 |  |  |  |
| 0 |  | 78,979 (21%) | 1,857 (22%) | 898 (25%) |
| 1 |  | 288,508 (79%) | 6,454 (78%) | 2,680 (75%) |
| Physical activity | 379,376 |  |  |  |
| 0 |  | 66,548 (18%) | 1,732 (21%) | 749 (21%) |
| 1 |  | 300,939 (82%) | 6,579 (79%) | 2,829 (79%) |
| Sedentary time | 379,376 |  |  |  |
| 0 |  | 98,619 (27%) | 2,506 (30%) | 1,155 (32%) |
| 1 |  | 268,868 (73%) | 5,805 (70%) | 2,423 (68%) |
| Sleep duration | 379,376 |  |  |  |
| 0 |  | 93,249 (25%) | 2,228 (27%) | 1,045 (29%) |
| 1 |  | 274,238 (75%) | 6,083 (73%) | 2,533 (71%) |
| Fruit & vegetable intake | 379,376 |  |  |  |
| 0 |  | 69,808 (19%) | 1,745 (21%) | 726 (20%) |
| 1 |  | 297,679 (81%) | 6,566 (79%) | 2,852 (80%) |
| Oily fish intake | 379,376 |  |  |  |
| 0 |  | 158,591 (43%) | 3,742 (45%) | 1,531 (43%) |
| 1 |  | 208,896 (57%) | 4,569 (55%) | 2,047 (57%) |
| Red meat intake | 379,376 |  |  |  |
| 0 |  | 32,600 (8.9%) | 769 (9.3%) | 332 (9.3%) |
| 1 |  | 334,887 (91%) | 7,542 (91%) | 3,246 (91%) |
| Processed meat intake | 379,376 |  |  |  |
| 0 |  | 113,419 (31%) | 2,694 (32%) | 1,195 (33%) |
| 1 |  | 254,068 (69%) | 5,617 (68%) | 2,383 (67%) |
| Social activity | 379,376 |  |  |  |
| 0 |  | 30,096 (8.2%) | 685 (8.2%) | 302 (8.4%) |
| 1 |  | 337,391 (92%) | 7,626 (92%) | 3,276 (92%) |
| Lifestyle score (LS) | 379,376 | 8 (7, 9) | 8 (7, 9) | 8 (7, 9) |
| Lifestyle status | 379,376 |  |  |  |
| Favorable |  | 138,221 (38%) | 2,753 (33%) | 1,145 (32%) |
| Intermediate |  | 167,549 (46%) | 3,880 (47%) | 1,648 (46%) |
| Unfavorable |  | 61,717 (17%) | 1,678 (20%) | 785 (22%) |
| LS metabolomic score (LS.mets) | 188,931 | 205.89 (205.55, 206.24) | 205.78 (205.45, 206.15) | 205.78 (205.43, 206.12) |
| Median (Q1, Q3); n (%) |  |  |  |  |

We implemented a three-step machine learning framework to identify metabolomic features associated with a healthy lifestyle using 327 plasma NMR biomarkers profiled by Nightingale Health (Supplementary Table 1). Specifically, participants who underwent NMR profiling at baseline recruitment were randomly divided into a 50% training dataset and a 50% optimization dataset, while the revisit cohort was reserved as an independent validation dataset. Repeated least absolute shrinkage and selection operator (LASSO) regression with 200 iterations was performed to identify stably selected NMR features in the training dataset. A total of 112 NMR features with a selection frequency exceeding 80% were carried forward to optimize their weights in the optimization dataset. Model performance was evaluated in the validation dataset, and no evidence of overfitting was observed (Methods).

The 112 healthy lifestyle-related metabolomic features spanned 13 metabolic groups, including amino acids, glycolysis-related metabolites, ketone bodies, fluid balance biomarkers, inflammatory biomarkers, fatty acids, other lipids, and six lipoprotein-related phenotypes (**Figure 2b**). Among these, 28 features were consistently selected across 200 iterations of LASSO regression (Supplementary Table 2) and were closely linked to cardiovascular health (e.g., HDL_size, M_LDL_TG, Omega_3_pct)[19, 20], energy metabolism (e.g., citrate, β-Hydroxybutyrate, branched-chain amino acids)[21, 22], and systemic inflammation (e.g., GlycA and Albumin)[23, 24].

Subsequently, we explored the correlations between a healthy lifestyle pattern and each of the 112 selected NMR features. Overall, 47 (42.0%) NMR features were classified as ’health-promoting’ due to their positive correlations with higher lifestyle scores. Typical examples included DHA_pct, omega_3_pct, PUFA_pct, PUFA_by_MUFA, and omega_3. In contrast, 53 (47.3%) NMR features were classified as ’health-compromising’ due to their negative correlations with lifestyle scores, such as omega_6_pct_PUFA and omega_6_by_omega_3 (Bonferroni-corrected *p*-values < 0.05) (**Figure 2c**). Specifically, diet-related factors—particularly oily fish, fruit and vegetable, and processed meat intake—along with alcohol consumption and smoking status, exhibited extensive metabolomic associations with the selected features. In contrast, sleep duration and social activity showed comparatively sparse associations (**Figure 2d**). Higher oily fish intake has been linked to a reduced risk of cardiovascular disease and mortality[25]. Consistent with previous findings[26], we observed that oily fish intake was strongly associated with higher levels of ’health-promoting’ omega-3 fatty acids (correlation coefficients: 0.09 for omega_3, 0.13 for omega_3_pct, and 0.13 for DHA_pct) and lower levels of ’health-compromising’ biomarkers (correlation coefficients: -0.10 for omega_6_pct_PUFA and -0.11 for omega_6_by_omega_3). These results underscore the significant benefits of including oily fish as part of a regular diet.

Due to the multicollinearity inherent in metabolomic data[27], we observed strong correlations among the 112 selected features (Supplementary Figure 1). To address this, we developed a healthy lifestyle-related metabolomic score (LS.mets) by assigning optimized weights to each feature, effectively accounting for the underlying correlation structure (Methods). This score provided a comprehensive measure of a participant’s lifestyle health based on their metabolomic profile and was carried forward into subsequent downstream analyses.

### Associations of lifestyle and related metabolomic profile with incident psoriasis risk

Lifestyle-related factors have been widely reported to be associated with the risk of psoriasis[10–12]. Here, we revisited these associations using a more comprehensive lifestyle score and further explored the association between a lifestyle-related metabolomic profile and incident psoriasis risk.

The Kaplan-Meier survival analysis (**Figure 3a**) revealed a strong link between lifestyle patterns—favorable, intermediate, and unfavorable—and psoriasis incidence over a median follow-up of 14.3 years. Notably, poorer lifestyle patterns were associated with a significantly higher risk of psoriasis (log-rank test *p* = 1.93E-19). To assess the robustness of these findings against potential confounding, we constructed four multivariate Cox proportional hazards models with varying sets of covariates (Methods). Across all models, individuals in the intermediate lifestyle group showed a modestly elevated risk of psoriasis compared with those in the favorable group, with hazard ratios (HRs) ranging from 1.14 to 1.15. Those in the unfavorable lifestyle group exhibited a substantially higher risk, with HRs between 1.38 and 1.44 (**Figure 3b**). A strong dose-response relationship was observed, with psoriasis risk increasing significantly as lifestyle patterns worsened (*p*-trend < 2.69E-10 in all models), consistent with prior evidence[10].

**Figure 3.**
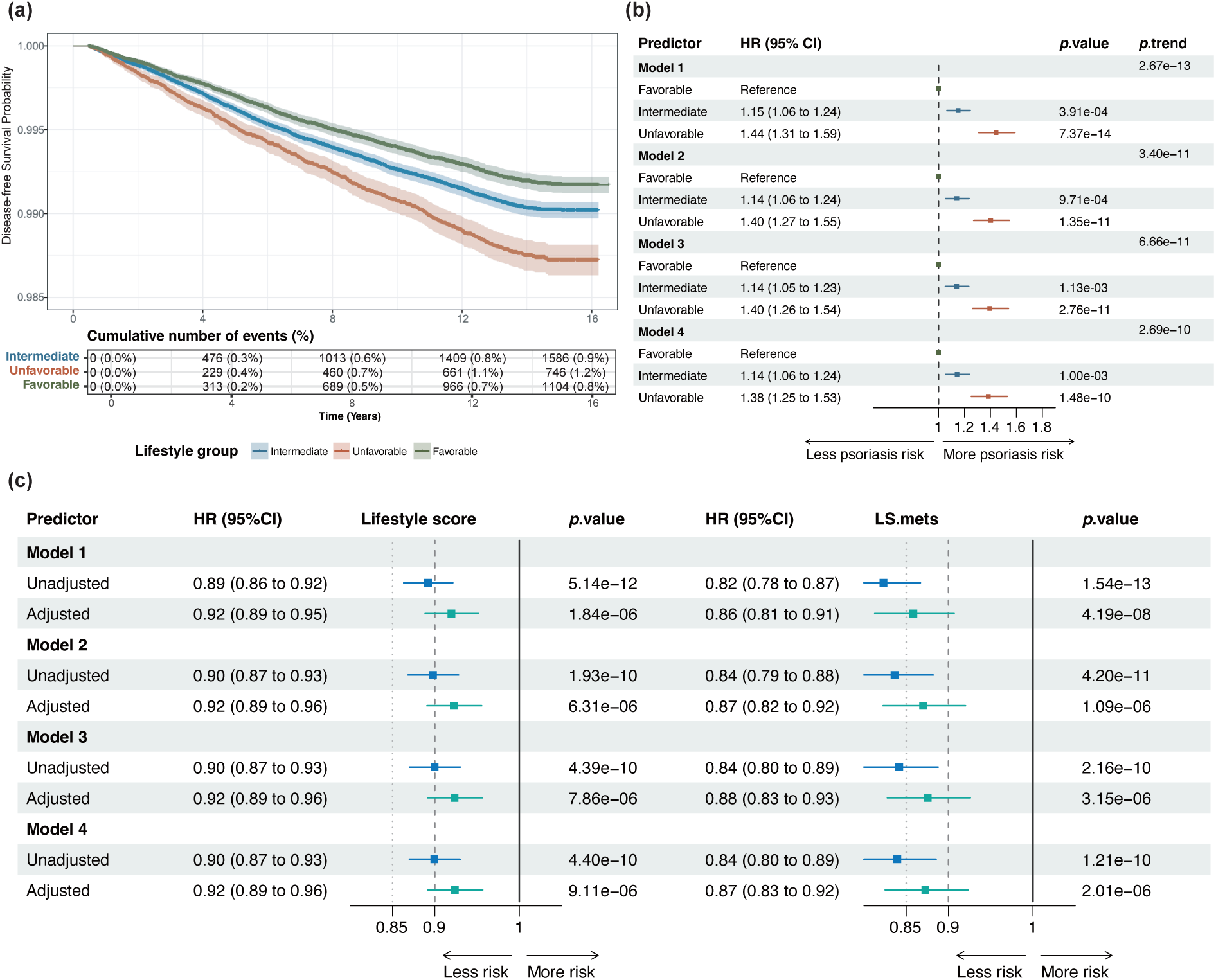
Associations of the lifestyle score (LS) and the lifestyle-related metabolomic score (LS.mets) with incident psoriasis risk. (a) Kaplan–Meier curves illustrating the cumulative incidence of psoriasis among participants categorized into unfavorable (*N*=62,382), intermediate (*N*=169,026), and favorable (*N*=139,272) lifestyle groups. The table below shows the number of events (n%, proportion) for each group at different time points. (b) Forest plot displaying hazard ratios (HRs) and 95% confidence intervals (CIs) for psoriasis across three lifestyle groups (*N*= 370,680), using the favorable group as the reference. Model 1 adjusts for age, sex, and body mass index (BMI). Model 2 additionally adjusts for ethnic background, living country, and index of multiple deprivation (IMD). Model 3 further adjusts for myocardial infarction, stroke, heart failure diagnosis, and cholesterol-lowering medication. Model 4 further adjusts for polygenic risk score (PRS) of psoriasis. The *p*-value for trend was calculated by treating the lifestyle group (ordered as favorable, intermediate, and unfavorable) as a continuous variable. (c) Hazard ratios for psoriasis risk associated with LS (per point) and LS.mets (per standard deviation) (*N*= 184,477). In unadjusted models, each score was evaluated as the individual predictor. In adjusted models, both LS and LS.mets were jointly fitted, thereby isolating their independent associations with psoriasis risk.

Our investigation into the association between the metabolomic profile and incident psoriasis risk revealed that both a higher lifestyle score and a higher LS.mets were significantly associated with a lower risk of psoriasis (**Figure 3c**). Specifically, each point increase in the lifestyle score corresponded to a 10-11% reduction in psoriasis risk (HR range: 0.89-0.90 across four models, all *p*-values < 4.40E-10), while each standard deviation increase in LS.mets was associated with a 16-18% risk reduction (HR range: 0.82-0.84 across four models, all *p*-values < 2.16E-10). Notably, LS.mets showed a stronger association than the lifestyle score, with consistenly lower HRs and more significant *p*-values, suggesting that the metabolomic profile may capture biological risk more precisely.

To assess their independent contributions, we jointly analyzed both predictors across four models. After mutual adjustment, the lifestyle score remained significant with HRs around 0.92 (*p*-values < 9.11E−06), while LS.mets retained a stronger association with HRs between 0.86 and 0.88 (*p*-values < 3.15E−06).

Stratified analyses by age (40-50, 50-60, and 60-70 years), sex (male and female), and polygenic risk of psoriasis (low, middle, and high) yielded similar results across different subgroups, supporting our primary observations (Supplementary Table 3-5). Since UK Biobank participants were middle-aged at recruitment, individuals with persistently unhealthy lifestyles may have developed psoriasis prior to their recruitment[28]. Restricting the analyses to post-recruitment incident cases could thus underestimate the true contributions of lifestyle and its metabolomic profile to psoriasis risk[29]. To address this potential bias, we conducted sensitivity analyses among prevalent cases (*N* = 11,053) to reassess associations of lifestyle and LS.mets with psoriasis (Methods). The results from sensitivity analyses remained largely consistent with those observed in the incident analyses, although the association of lifestyle suffered greater attenuation after adjusting for LS.mets (Supplementary Figure 2).

Findings above highlighted that lifestyle and related metabolomic profile contributed independently to psoriasis risk. However, the attenuated association between the lifestyle score and psoriasis after adjusting for LS.mets suggested that part of the effect of lifestyle may be mediated through metabolomic pathways, underscoring the potential biological relevance of metabolism in capturing the downstream effects of lifestyle on disease risk.

### Mediating effects of the metabolomic profile in “lifestyle-psoriasis” association

To explore this further, we conducted mediation analyses using a natural effect framework to explicitly and systematically assess how LS.mets and metabolomic features mediate the association between lifestyle and psoriasis risk (Methods). The natural indirect effects (NIE)—representing the effect of lifestyle mediated specifically through LS.mets—were consistently significant across all four models (OR=0.96–0.97, *p*-values < 5E-04) (**Figure 4a**). These NIE estimates indicated that lifestyle-related metabolomic changes substantially mediated the association, accounting for approximately 46.1% (Model 1), 40.6% (Model 2), 40.2% (Model 3), and 37.5% (Model 4) of the total effect of the lifestyle. After extracting this metabolomic-mediated component, the natural direct effects (NDE), representing the remaining direct associations between lifestyle and psoriasis, remained statistically significant but attenuated (OR=0.95–0.96, *p*-values < 0.05).

**Figure 4.**
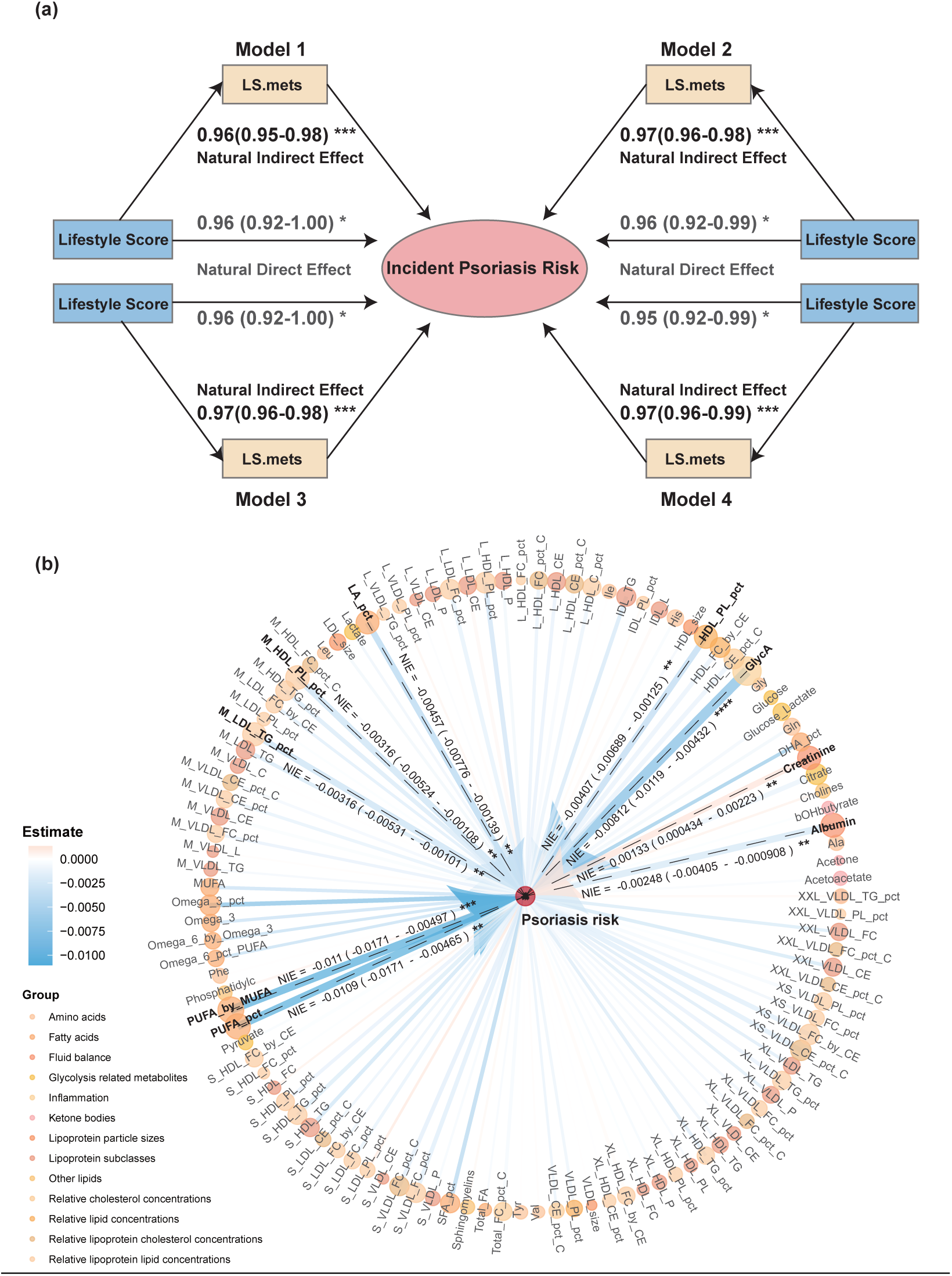
Metabolomic features mediate the association between lifestyle and psoriasis. (a) Path diagrams showing the natural indirect effect (NIE) and natural direct effect (NDE) of the lifestyle score (LS) on psoriasis via the lifestyle-related metabolomic score (LS.mets) across four models (*N*= 184,477). Both the NIE and NDE, with their 95% confidence intervals, were obtained by exponentiating the coefficients from the natural effect model (***, *p*-value<5E-04; **, *p*-value<5E-03; *, *p*-value<5E-02). (b) Circular plot illustrating the mediating effects of each NMR feature on psoriasis risk, estimated with Model 4 (*N*=184,477). The 112 NMR features are arranged around the perimeter and color-coded by metabolic category, while the center node represents psoriasis as the outcome. The color gradient indicates NIE directly obtained from the coefficient in the natural effect model. The edge width and node size are proportional to the negative logarithm of the *p*-value, reflecting the strength of mediation. Nine mediators passing the Bonferroni-corrected significance threshold are highlighted in bold (****, *p*-value<5E-05; ***, *p*-value<5E-04; **, *p*-value<5E-03).

To pinpoint specific biomarkers underlying this mediation effect, we further conducted pairwise mediation analyses with each of the 112 NMR features individually tested as mediators (Methods). Among these, nine biomarkers exhibited consistently significant NIE across all four models (Bonferroni-corrected *p*-values < 0.05) (**Figure 4b**). These mediators spanned multiple metabolic pathways including inflammation (GlycA and albumin), polyunsaturated fatty acid metabolism (PUFA/MUFA, PUFA_pct, and LA_pct), lipoprotein particle properties (M_HDL_PL_pct, HDL_PL_pct, M_LDL_TG_pct), and renal function (creatinine) (Supplementary Table 6).

Given the multicollinearity among NMR features, pairwise mediation analyses may inadequately address intercorrelations among candidate mediators, limiting their ability to pinpoint independent mediators[30]. To overcome this limitation, we employed a high-dimensional mediation analysis framework specifically designed for omics data, enabling robust identification of independent mediators despite strong correlations among metabolomic features (Methods). Using this approach, four NMR features were consistently identified as independent and significant mediators after correcting for multiple-testing: GlycA (*p*-value = 8.86E-11 in Model 4), PUFA/MUFA (*p*-value = 1.47E-09 in Model 4), VLDL_size (*p*-value = 1.07E-06 in Model 4), and creatinine (*p*-value = 8.14E-5 in Model 4) (Supplementary Table 7). Importantly, three of these biomarkers— GlycA, PUFA/MUFA, and creatinine—were previously highlighted as significant mediators in pairwise analyses. Their consistent identification in both analytical frameworks provided robust statistical support for their involvement in biological pathways linking lifestyle to psoriasis.

### Shared genetic architectures between psoriasis and independent metabolomic mediators

To explore underlying biological mechanisms by which GlycA, PUFA/MUFA, and creatinine mediate the relationship between the lifestyle and psoriasis, we examined their GWAS summary statistics and identified shared genetic architectures between these biomarkers and psoriasis, suggesting common biological pathways that may contribute to their associations (Methods).

GWAS signals for GlycA substantially overlapped with psoriasis-associated genetic loci within the major histocompatibility complex (MHC) region (shared locus: chr6: 30,803,497 - 33,426,466) (**Figure 5a**). This genomic region is characterized by a highly complex genetic architecture and is crucial in both adaptive and innate immune responses[31]. The overlapping signals mapped to MHC class I and class II genes— including HLA-C, HLA-DRA, HLA-DQA1, HLA-DQA2, and HLA-DQB1—as well as to genes involved in Notch signaling (NOTCH4) and collagen fibril organization (COL11A2). A near-perfect proxy for the major psoriasis risk genotype HLA-C*06:02, rs4406273[32], was also associated with GlycA (*p*-value=1.10E-05). The convergence of these genetic signals in the MHC region suggests that GlycA, as a systemic inflammation biomarker, may mediate psoriasis risk through immune mechanisms related to antigen presentation[33].

**Figure 5.**
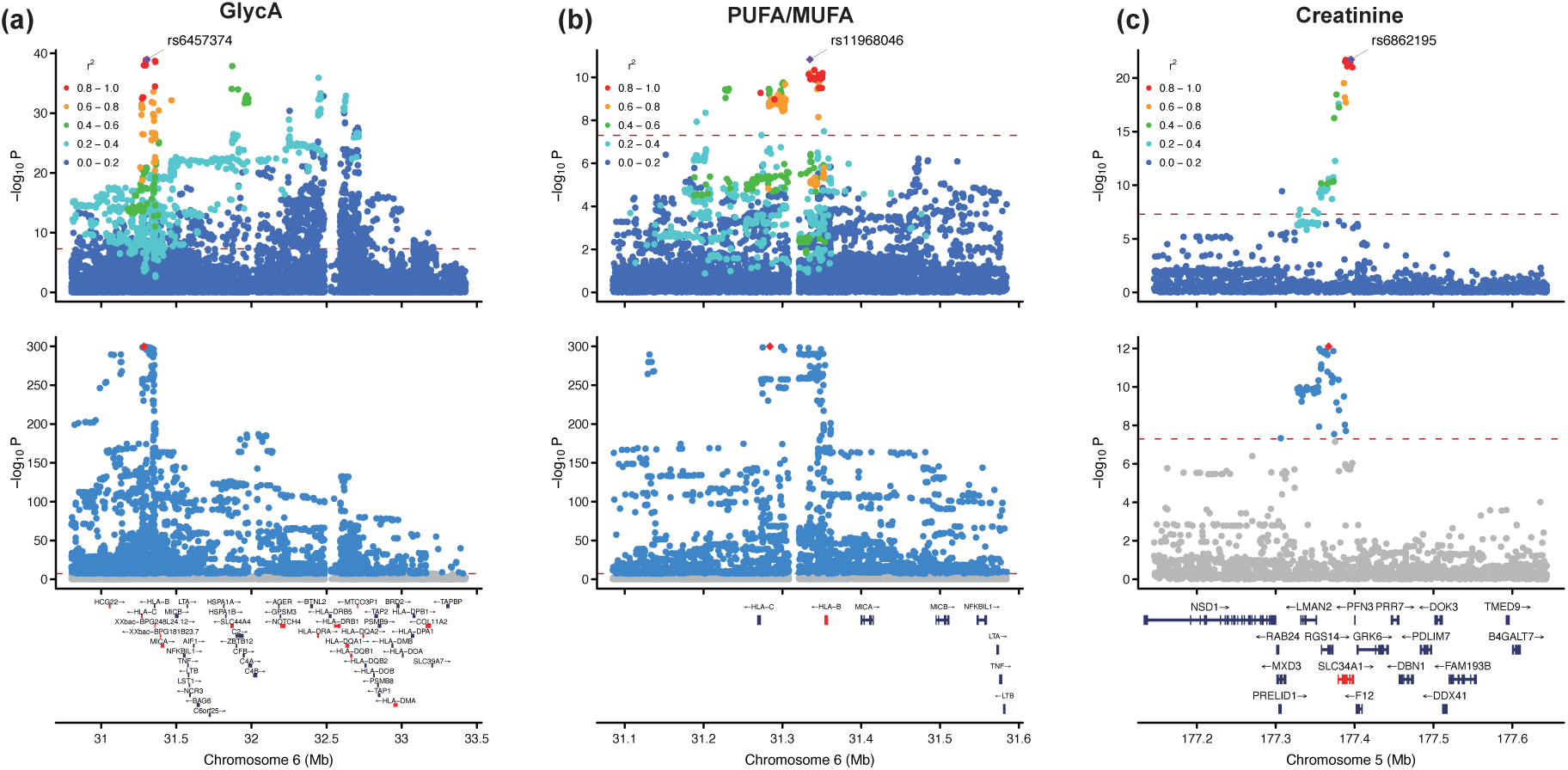
Shared genetic architectures of GlycA, PUFA/MUFA, and creatinine with psoriasis. (a) LocusZoom plots showing overlapping GWAS signals of GlycA (top panel) and psoriasis (bottom panel) in the locus: chr6:30,803,497-33,426,466 (hg38 build). (b) LocusZoom plots showing overlapping GWAS signals of PUFA/MUFA (top panel) and psoriasis (bottom panel) in the locus: chr6: 31,084,865-31,584,865 (hg38 build). (c) LocusZoom plots showing overlapping GWAS signals of creatinine (top panel) and psoriasis (bottom panel) in the locus: chr5:177,145,511-177,645,511 (hg38 build). Sentinel variants from the NMR biomarker GWAS are labeled, and the nearest genes for independent SNPs (after clumping the NMR GWAS summary statistics) are highlighted in red. The lead SNP in the psoriasis GWAS is also highlighted in red. The red dashed line denotes the genome-wide significance threshold (*p*-value of 5E-08). Linkage disequilibrium (LD) r² values with the sentinel variant were calculated using whole-genome sequencing data from 95,372 participants (see Methods).

GWAS signals for PUFA/MUFA also substantially overlapped with psoriasis-associated genetic loci within the MHC region and mapped to genes including HLA-B and HLA-C (shared locus: chr6: 31,084,865 - 31,584,865) (**Figure 5b**). Dietary supplementation with polyunsaturated fatty acids has been reported to have immunosuppressive effects through the human leukocyte antigen (HLA) class I pathway[34]. Specifically, the incorporation of PUFAs lowers the susceptibility of the target cell to lysis by effector T cells and diminishes the formation of antigen-presenting cell-T cell conjugation[34]. The convergence of genetic signals at HLA class I loci and the immunomodulatory effects of PUFAs suggests that altered lipid metabolism may influence psoriasis risk by reducing antigen presentation and T cell activation, highlighting a potential shared immunoregulatory pathway.

GWAS signals for creatinine overlapped with psoriasis-associated genetic loci on chromosome 5, mapping to SLC34A1 (shared locus: chr5: 177,145,511-177,645,511) (**Figure 5c**). SLC34A1 encodes the sodium-phosphate cotransporter NaPi-IIa, a key regulator of phosphate reabsorption in the renal proximal tubule[35]. Mutations in this gene have been linked to chronic kidney disease[36], a common comorbidity observed in individuals with psoriasis. The convergence of genetic signals at this locus suggests that altered phosphate transport and renal function might provide a link between creatinine levels and psoriasis.

Additional overlapping genetic signals between the three biomarkers and psoriasis across other genomic regions were presented in Supplementary Figure 3, offering further insights into the potential biological mechanisms underlying their associations with psoriasis.

### Associations of lifestyle and related metabolomic profile with eight clinical prognostic outcomes

To extend our findings on incident psoriasis risk, we examined whether a healthy lifestyle and its related metabolomic profile were also associated with potential clinical outcomes among psoriasis patients, which could provide guidance on how to prevent subsequent comorbidities for them.

We examined eight clinical prognostic outcomes, including all-cause mortality, psoriatic arthritis—the most typical complication of psoriasis[37]—and six common comorbidities (cardiovascular diseases, obesity-related conditions, gastrointestinal diseases, multiple cancers, mood disorders, and chronic kidney disease) that psoriasis patients are more likely to develop compared to those psoriasis-free participants (Supplementary Figure 4).

Across all eight clinical outcomes, a healthier lifestyle score and a more favorable LS.mets—particularly higher PUFA/MUFA and lower GlycA levels—were consistently associated with reduced prognostic risk among prevalent psoriasis cases (**Figure 6a**). Specifically, higher LS.mets were significantly associated with lower prognostic risk for all-cause mortality (HR=0.728), psoriatic arthritis (HR=0.787), CVDs (HR=0.866), obesity-related conditions (HR=0.825), mood disorders (HR=0.731), multiple cancers (HR=0.904), chronic kidney disease (CKD, HR=0.841), and gastrointestinal diseases (HR=0.742) (Bonferroni-corrected *p*-values < 0.05 and nominal *p*-values < 0.05 for multiple cancers and CKD). Similar but slightly attenuated associations were also observed for the lifestyle score (Supplementary Table 8).

**Figure 6.**
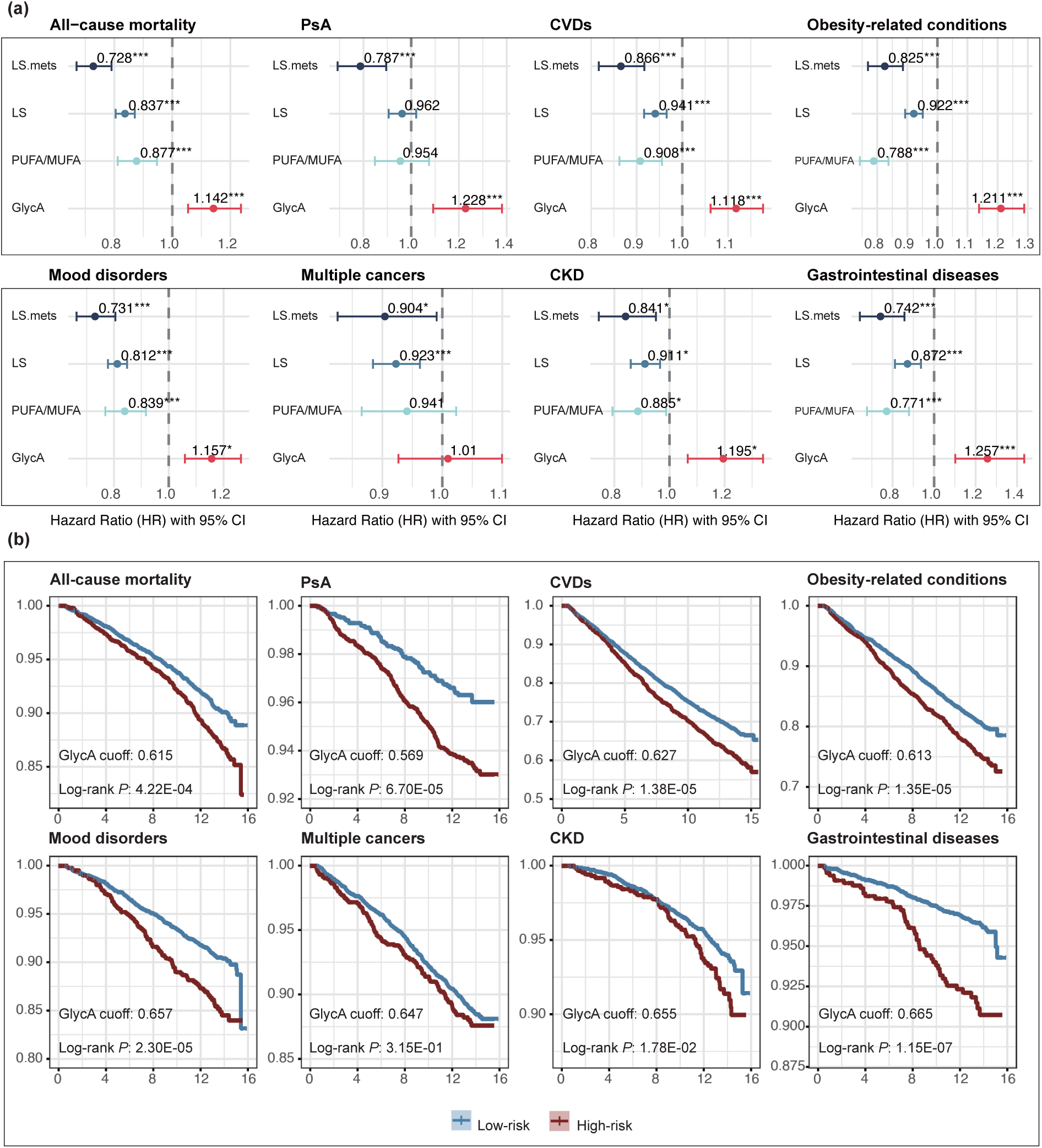
Associations of the lifestyle score (LS), metabolomic score (LS.mets), and key metabolomic mediators with eight prognostic outcomes in prevalent psoriasis patients (*N*=15,978). (a) Forest plots displaying hazard ratios (HRs) and 95% confidence intervals (CIs) for LS.mets (scaled), LS, PUFA/MUFA (scaled), and GlycA (scaled) associated with eight clinical outcomes: all-cause mortality, psoriatic arthritis (PsA), cardiovascular diseases (CVDs), obesity-related conditions, mood disorders, multiple cancers, chronic kidney disease (CKD), and gastrointestinal diseases (***, *p*-value<1.25E-03 defined by Bonferroni-correction; *, *p*-value<0.05). (b) Kaplan-Meier curves for the same outcomes, stratified by low-risk versus high-risk groups based on optimal GlycA cutoffs (see Methods). The x-axis represents follow-up time, and the y-axis indicates survival or event-free probability. Differences between groups were compared using the log-rank test.

Among three metabolomic mediators, PUFA/MUFA levels were inversely associated with all prognostic outcomes, reaching statistical significance in six of eight conditions, with the strongest protective effect observed for obesity-related conditions (HR=0.788, *p*-value=1.12E-14). In contrast, GlycA as an inflammatory biomarker was positively associated with increased risk across nearly all prognostic outcomes except for multiple cancers. Notably, higher GlycA levels were significantly associated with higher risk of gastrointestinal diseases (HR=1.26, *p*-value=6.60E-04), psoriatic arthritis (HR=1.23, *p*-value=6.15E-04), obesity-related conditions (HR=1.21, *p*-value=1.35E-09), and CVD (HR=1.12, *p*-value=2.23E-05), suggesting its potential clinical value as a poor prognosis predictor in psoriasis patients. Higher creatinine levels were strongly associated with higher CKD risk (HR=2.01, *p*-value=8.09E-65) but showed no significant associations with other outcomes after Bonferroni correction (Supplementary Table 9).

### GlycA as a potential prognostic biomarker

To further assess the predictive performance of lifestyle score, LS.mets, PUFA/MUFA, and GlycA levels across eight prognostic outcomes, we performed a series of precision-recall curve analyses (Methods). We noticed that GlycA consistently demonstrated the highest predictive value across all outcomes except for multiple cancers (Supplementary Figure 5). Thus, we prioritized GlycA as a candidate prognostic biomarker for psoriasis patients and subsequently evaluated comorbidity incidence and survival outcomes across GlycA risk groups (Methods).

Kaplan-Meier analysis revealed that psoriasis patients in the high-GlycA group had significantly worse prognoses across multiple clinical outcomes, with consistently higher mortality rate and comorbidity incidence compared to the low-GlycA group (**Figure 6b**). As there is currently no recognized biomarker for psoriasis prognosis[38], these findings highlight potential clinical value of GlycA as a novel prognostic indicator for psoriasis patients.

## Discussion

Healthy lifestyle patterns have long been associated with a reduced risk of psoriasis[10, 12], yet the underlying mechanisms and pathways remain largely unknown. Here, we demonstrate that a healthy lifestyle-related metabolomic profile largely mediates the relationship between healthy behaviors and psoriasis risk, highlighting key metabolomic mediators and suggesting potential biological mechanisms. We further extend these associations to multiple clinical prognostic outcomes in psoriasis patients, offering comprehensive insights that may inform early prevention and personalized interventions for psoriasis and subsequent comorbidities.

We constructed a comprehensive measure indicating a healthy lifestyle pattern encompassing multiple factors, including smoking status, alcohol consumption, physical activity, sedentary time, sleep duration, fruit and vegetable intake, oily fish intake, red meat intake, processed meat intake, and social activity. Leveraging extensive metabolomic data of 327 NMR biomarkers from 275,226 UK Biobank participants, we have expanded the healthy lifestyle-related metabolomic profile to the largest scale so far. Through a three-step machine learning framework, we identified 112 lifestyle-associated NMR features over 200 iterations, from which we derived a novel metabolomic score that captures the biological imprint of a healthy lifestyle. These NMR features span various metabolic pathways and biological functions, such as glycolysis, lipid metabolism, fluid balance, and inflammation, underscoring the wide-ranging impacts of external lifestyle exposures on intrinsic plasma metabolome[39].

Our investigation into the association between lifestyle and incidence psoriasis risk supports previous studies. We found that both a healthy lifestyle score and a favorable metabolomic score were independently and jointly associated with reduced psoriasis risk. Notably, the metabolomic score exhibited a stronger association than the lifestyle score. Several factors may explain this enhanced association. First, metabolomic changes act as intrinsic messengers that integrate environmental signals, serving as a more direct link with disease mechanisms[40]. Second, subtle lifestyle changes can trigger biological cascades that amplify metabolic alterations[41], thereby influencing key pathways in psoriasis pathogenesis. Third, metabolomic biomarkers offer greater sensitivity and specificity to early changes in disease progression, providing a more precise and dynamic snapshot of health status[42]. Finally, the metabolomic profile may capture not only the direct effects of lifestyle factors but also higher-level interactions between lifestyle and genetic components[43], yielding a more comprehensive risk assessment. These characteristics underscore the potential of the plasma metabolome as a more effective disease risk predictor.

Recently, growing evidence has emphasized the pivotal role of metabolism in psoriasis pathogenesis, particularly in keratinocytes[44]. Our systematic mediation analyses reveal that the metabolomic profile mediates approximately 37.5–46.1% of the total effect of lifestyle on psoriasis. Pairwise and multivariate mediation analyses further identify three independent metabolomic biomarkers—GlycA, PUFA/MUFA, and creatinine—as key mediators, underscoring the involvement of systemic inflammation, polyunsaturated fatty acid metabolism, and creatinine-related pathways (e.g., energy metabolism and renal function) in psoriasis pathology.

Through investigations into the genetic architectures for GlycA, PUFA/MUFA ratio, creatinine, and psoriasis, we identified shared genomic regions with overlapping association signals, offering insights into how these biomarkers may mediate psoriasis risk. For both GlycA and PUFA/MUFA, the convergent signals mapped to the MHC region, particularly HLA genes. Notably, GlycA levels demonstrate a dose-response with psoriasis severity[45], suggesting that T-cell activation and antigen presentation may underlie its association with psoriasis[46]. Because multiple acute-phase proteins (e.g., α1-acid glycoprotein, haptoglobin, α1-antitrypsin, α1-antichymotrypsin, and transferrin) contribute to GlycA signals during NMR profiling[47], their specific roles remain to be elucidated.

Similarly, the shared signals for PUFA/MUFA and psoriasis localized to HLA-B and HLA-C. PUFAs can reduce HLA class I surface expression and ER–Golgi trafficking, thereby lowering target-cell susceptibility to cytotoxic T lymphocytes and impairing antigen-presenting cell–T cell conjugation[34]. This aligns with the complex interplay among CD4⁺, CD8⁺ T cells, and cross-presenting dendritic cells in psoriatic lesions[48], providing a plausible mechanism for PUFAs’ protective effect. We advocate that a balanced PUFA/MUFA ratio of 1:1, as recommended by the American Heart Association[49], may confer anti-inflammatory benefits and help prevent psoriasis.

By contrast, the biological link between creatinine and psoriasis is less established, although psoriasis patients often exhibit elevated creatinine[50] and a higher risk of CKD[51]. Shared genetic signals mapped to SLC34A1, which encodes the renal sodium-phosphate cotransporter NaPi-IIa[35]. This transporter is crucial for sodium and phosphate reabsorption, phosphate homeostasis, and regulation of serum 1,25-(OH)₂D₃ [52]. These findings suggest that disruptions in renal phosphate handling and vitamin D metabolism may represent a novel metabolic-renal pathway contributing to psoriasis risk, warranting further exploration.

Subsequent comorbidities following a psoriasis diagnosis can impose a substantial medical cost burden[53], significantly deteriorate patient’s health conditions[54], and shorten their life expectancy[4], emphasizing the need for early preventive measures. By extending our analysis from incident to prevalent psoriasis cases, we found that maintaining a healthy lifestyle and a favorable metabolomic profile markedly lowered the risk of multiple clinical prognostic outcomes. Given the high healthcare expenses linked to psoriasis-related comorbidities—such as CVDs, obesity-related conditions, psoriatic arthritis, and elevated mortality—promoting healthy lifestyle interventions and screening for metabolomic indicators may help reduce healthcare costs and improve patient quality of life.

Our study also highlights GlycA as a potential prognostic biomarker and risk predictor for psoriasis, particularly in the current absence of any widely accepted marker of disease progression [38]. Future research could refine GlycA by pinpointing its most predictive components, optimizing cutoff values for sensitivity and specificity, and conducting large-scale prospective validations to establish its clinical utility.

Our study has several limitations. First, participants in the UK Biobank tend to be healthier and less deprived than the general UK population[55], which may underestimate the impact of an unhealthy lifestyle and associated metabolomic profile on psoriasis risk. Second, because the cohort was recruited mainly between ages 40 and 60, the relationship between a healthy lifestyle, associated metabolomic profile, and early-onset psoriasis—often peaking in the 20s and 30s[28] and carrying a stronger genetic predisposition[56]—remains underexplored. Although our sensitivity analyses included these early-onset cases and accounted for polygenic risk, further research in younger prospective cohorts would add valuable insight. Third, our study population is predominantly of European ancestry, and while psoriasis prevalence is high among Europeans[3], extending the analysis to other ancestries would help bridge current health disparities. Fourth, although shared genetic architectures implicated potential mechanisms underlying the mediating effects of three metabolomic biomarkers, the specific biological pathways remain elusive and warrant deeper investigation. Lastly, due to limited revisit data, our cross-sectional design does not capture dynamic changes in metabolomic profiles over time. A longitudinal approach would present more nuanced and personalized associations between metabolomic changes reflective of lifestyle patterns and psoriasis risk[57].

To conclude, our study establishes a comprehensive metabolomic profile of health lifestyle behavior that not only mediates the association with psoriasis risk but also serves as a predictor of broader clinical outcomes. This integrative profile captures the systemic biological imprint of lifestyle choices, offering a promising avenue for early risk stratification, targeted prevention, and precision health strategies in both psoriasis and multiple comorbidities.

## Methods

### Study populations

The UK Biobank project was approved by the National Information Governance Board for Health and Social Care and the North West Multicentre Research Ethnical Committee (11/NW/0382)[18]. 502,129 participants recruited between 2006 and 2010 and providing written informed consent were included in our analysis. This research was conducted under application ID 548711.

### Construction of a healthy lifestyle score

Ten healthy lifestyle components were defined and assessed based on previous literatures[58–60] and UK national health service guidelines (https://www.nhs.uk/live-well/). They included: (1) smoking status (1 point for past or never smoker; 0 point for current smoker); (2) alcohol consumption (1 point for ≤ 4 times a week; 0 point for daily or almost daily); (3) physical activity(1 point for ≥ 150 minutes/week moderate physical activity or ≥ 75 minutes/week vigorous physical activity; 0 point for < 75 minutes/week vigorous physical activity); (4) sedentary time (1 point for < 4 hours/day; 0 point for ≥ 4 hours/day); (5) sleep duration (1 point for 7 to 9 hours/day; 0 point for less than 7 hours or more than 9 hours per day); (6) fruit and vegetable intake (1 point for ≥ 400 grams/day; 0 point for < 400 grams/day); (7) oily fish intake (1 point for ≥ 1 portion/week; 0 point for < 1 portion/week); (8) red meat intake (1 point for ≤ 3 portions/week; 0 point for > 3 portions/week); (9) processed meat intake (1 point for ≤ 1 portion/week; 0 point for > 1 portion/week); (10) social activity (1 point for friends and family visits more than once a month; 0 point for friends and family visits less than once a month). Details on data process using relevant UK Biobank fields were provided in <u>Supplementary Note 1</u>. One point was assigned for each healthy lifestyle criterion met, contributing to the overall lifestyle score (ranging from 0 to 10). Participants were further divided into three groups based on their lifestyle score as: favorable (9 - 10), intermediate (7 - 8), and unfavorable (0 - 6).

### Quality control of nuclear magnetic resonance (NMR) metabolomic data

This study used NMR metabolomic data (Phase 2 release version) covering 251 raw metabolomic biomarkers from 275,226 EDTA plasma samples. Metabolic biomarkers were measured using a high-throughput NMR-based profiling platform developed by Nightingale Health Ltd. Details about plasma sample preparation and biomarker profiling have been described elsewhere[61]. The pipeline of quality control and computation for additional biomarker ratios followed the steps below[62]:

#### (1) Data extraction

We extracted raw NMR biomarker data from both baseline assessments and the first repeat assessments. Quality control flags for each biomarker, NMR sample processing flags and quality control flags for each sample were also extracted.

#### (2) Removal of technical variation

Algorithms for removing technical variation sequentially adjusted the time between sample preparation and sample measurement, the systematic differences between rows and columns on the 96-well shipment plates, and the drift over time within each of the six spectrometers. Absolute concentrations were log-transformed before the removal of technical variation and regression residuals after the sequential adjustments were transformed back to absolute concentrations. Outliers of non-biological origins were identified and set to missing. Further details were provided and discussed by Ritchie *et al*[62].

#### (3) Computation of derived biomarkers and ratios after adjusting for biological variation

After removing technical variations, non-derived biomarkers were further adjusted for unwanted biological variations, including age, sex, and body mass index, using robust linear regression. Biomarkers were log-transformed prior to adjustment and transformed back to absolute concentrations after removing biological variations. Composite biomarkers were recomputed and 76 additional biomarker ratios not available in the original metabolomic data but with potential biological implications were derived.

Finally, a total of 327 NMR biomarkers (109 in absolute concentrations, 61 composite biomarkers, 81 biomarker ratios and 76 additionally derived biomarker ratios) were included for downstream analysis (Supplementary Table 1).

### Lifestyle-related metabolomic profiles

To identify healthy lifestyle-related metabolomic features and construct an overall metabolomic score, we followed a machine learning framework including feature selection, weight optimization, and independent validation[63]. Individuals with complete NMR data from the baseline assessment (*N* = 188,931) were randomly divided into a training dataset (50%, *N* = 94,467) for feature selection and an optimization dataset for hyperparameter tuning and weight optimization (50%, *N* = 94,464). Individuals with NMR data from the first revisit assessment (*N* = 12,917) were included in the out-of-sample validation dataset. NMR data for 327 biomarkers were log1p-transformed before model training. When performing feature selection in the training dataset, we used least absolute shrinkage and selection operator (LASSO) regression with the overall lifestyle score as the outcome of interest and the 327 NMR biomarkers as predictors. To maximize the stability and reproducibility of selected features, we resampled 200 random subsets, each comprising 50% of the training dataset. In each subset, we performed 10-fold cross-validation with five repeats and used a grid search to identify the model with the lowest root mean square error (RMSE). Across the 200 iterations, we identified varying combinations of NMR features and ranked them by their selection frequency. NMR features selected in more than 80% of iterations (> 160 times in total) were defined as “healthy lifestyle-related”. This process yielded 112 stable NMR features, which were carried forward for further analysis.

To optimize the weight of each NMR feature, we performed elastic net regression in the optimization dataset with the overall lifestyle score as the outcome and the 112 selected features as predictors. A grid search was conducted over a range of hyperparameters, including α (mixing parameter, ranging from 0 to 1 in increments of 0.05) and 11 (regularization parameter, ranging from 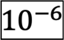 to 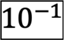 on a logarithmic scale). We employed 10-fold cross-validation with 10 repeats, and the optimal model, determined by the lowest RMSE, was selected with the best-tuning hyperparameters identified as α = 0.55 and λ = 0.001. Finally, the coefficients of the optimized model were extracted as the weights for each NMR feature (Supplementary Table 2). To avoid overfitting, we evaluated the optimized model’s performance in the validation dataset. The RMSE values were comparable between the optimization (RMSE = 1.35) and validation (RMSE = 1.31) datasets, indicating no evidence of overfitting.

The lifestyle-related metabolomic scores (LS.mets) were calculated for the entire cohort by computing a weighted sum of the selected NMR features, using coefficients from the optimized elastic net model. The Pearson’s correlation between the lifestyle score and LS.mets was 0.37 in the baseline assessment and 0.36 in the revisit assessment, demonstrating consistency and minimal overfitting.

### Effective number of metabolomic NMR features

To determine the effective number of independent metabolomic biomarkers, we followed an eigendecomposition method[64]. First, we calculated the covariance matrix of the 112 selected NMR features. Next, eigen decomposition was performed on the covariance matrix to extract its eigenvalues. Using these eigenvalues, we computed the effective number of independent metabolomic biomarkers using the formula:

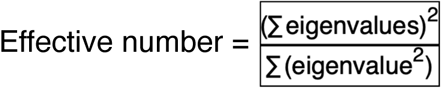

This yielded an effective number of 6.037, reflecting the independent dimensionality of the metabolomic data after accounting for its correlation structure.

### Multivariate Cox proportional hazards regression

To explore the associations of lifestyle score and LS.mets with incident psoriasis risk, we excluded prevalent cases at baseline (*N* = 11,053). We further excluded incident cases diagnosed within the first 6 months after baseline assessment (*N* = 211) to mitigate reverse causality. This 6-month exclusion window reduced bias from delays between symptom onset and formal diagnosis, reduced misclassification of prevalent cases, and ensured a temporal relationship between exposures and the risk of psoriasis[65]. After exclusions, the incident psoriasis cohort comprised 4,714 incident cases and 486,151 psoriasis-free controls. We fitted four multivariate Cox proportional hazards models with different sets of covariates to evaluate robustness to confounding. The baseline date was defined as the date each participant attended the assessment center (UK Biobank field ID: 53). The end of follow-up was set as the latest psoriasis diagnosis date (UK Biobank field ID: 131742; 2023-07-01). Participants who died during follow-up were censored at their date of death (UK Biobank field ID: 40000). For each participant, observation time was calculated from the baseline date to the earliest of three events: psoriasis diagnosis, death, or end of follow-up. Four multivariate Cox proportional hazards models were fitted using the formula:

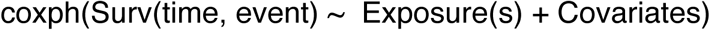

Model 1 adjusted for basic clinical factor including age, sex, and body mass index (BMI). Model 2 further adjusted for socio-demographic factors including ethnic background, living country, and Index of Multiple Deprivation (IMD, a composite measure of seven subdomains: housing, crime, education, employment, health, income, and living environment). Model 3 additionally adjusted for cardiovascular disease (CVD) burden and mediation[66], including stroke, heart failure, myocardial infarction, and cholesterol-lowering medication. Model 4 further adjusted for genetic risk using a psoriasis polygenic risk score (PRS). Exposures included lifestyle status as a categorical variable (favorable, intermediate, and unfavorable), lifestyle score as a continuous variable, and scaled LS.mets as a continuous variable, respectively. The lifestyle score and scaled LS.mets were further jointly fitted to explore their independent associations with incident psoriasis risk.

### Mediation analysis

Mediation analysis using a natural effect model was conducted to quantify the mediating effects of metabolomic profiles on the association between lifestyle score and incident psoriasis risk. Natural effect models, based on a counterfactual framework, decompose the total causal effect into natural direct effect (NDE) and natural indirect effect (NIE). Unlike traditional mediation strategies, this approach provides more interpretable results across different data distributions and effect scales[67].

First, we assessed whether LS.mets (mediator, ***M***) explained part of the relationship between lifestyle score (exposure, ***X***) and incident psoriasis risk (outcome, ***Y***). We fitted four natural effect models, adjusting for the same covariates (***C***) as in models 1-4 mentioned above. The mediator model, which predicts LS.mets based on lifestyle score and covariates, was formulated as:

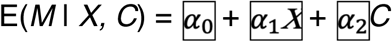

Using regression weights derived from this model, we fitted the natural effect model to decompose the total effect of lifestyle score on psoriasis risk into two parts[68]:

(1) Natural direct effect (NDE): the effect of lifestyle score on psoriasis risk if LS.mets were fixed at its natural value (i.e., the value it would take under a given lifestyle score).
(2) Natural indirect effect (NIE): the effect of lifestyle score on psoriasis risk that operates through LS.mets, capturing how changes in lifestyle score influence LS.mets, which in turn influence psoriasis risk.

The natural effect model was specified as:

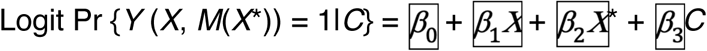

Here, *X* represents the actual lifestyle score, and *X*\* represents a hypothetical and counterfactual value of the lifestyle score. A significant NIE indicates that LS.mets mediates the relationship between lifestyle score and psoriasis risk. To quantify this mediation, we computed the mediation proportion (MP) using the following formula:

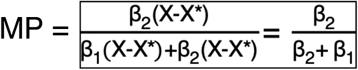

Additionally, we evaluated the potential mediating effect of each selected NMR feature using the natural effect model described above. This resulted in 112 pairwise mediation analysis, with each NMR feature treated as the mediator respectively. To account for multiple testing, we applied a Bonferroni-corrected p-value threshold of 0.05/6 (based on the effective number of features) to determine statistical significance.

Due to multicollinearity among NMR features, pairwise mediation analysis could not account for correlations among potential mediators and was suboptimal for identifying independent mediators. To address this issue, we used a high-dimensional mediation analysis framework called “HIMA”, which provides robust mediation effect estimates while controlling for type-I error[30]. The matrix of 112 NMR features was imported as a joint mediator matrix, with the type specified as “gaussian”. We chose the algorithm of high-dimensional mediation-based Cox model which involves three steps: mediator screening, de-biased LASSO estimation, and a joint significance test[69]. A false discovery rate (FDR) threshold of 0.05 was applied using Benjamini-Hochberg correction for multiple testing to report independent mediators.

### Identification of overlapped genetic architectures

GlycA, PUFA/MUFA, and creatinine were identified as independent mediators among 112 NMR biomarkers in both pairwise mediation analysis and the multivariate HIMA framework. To shed light on the potential mechanisms underlying their mediation of psoriasis risk, we investigated the shared genetic architectures of these three biomarkers and psoriasis. The genome-wide association study (GWAS) summary statistics for psoriasis were obtained from the FinnGen study[70] (R12 release: November 4, 2024), comprising 12,760 cases and 482,181 controls. For the three NMR biomarkers (GlycA, PUFA/MUFA, and creatinine), GWAS summary statistics were sourced from our previous publication, which utilized whole-genome sequencing (WGS) data from 95,372 UK Biobank participants who underwent metabolomic profiling[71]. To identify independent genetic loci, we performed clumping for NMR GWAS summary statistics using PLINK 2.0 alpha. The clumping parameters included a physical distance radius of 500 kilobases (kb), an LD threshold of 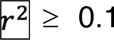, and a genome-wide significance threshold of 5E-08 for identifying index SNPs. Genomic loci were defined as ± 250 kb around each index SNP. Overlapping genomic loci were iteratively merged to ensure a set of independent loci for downstream analysis. For each NMR biomarker, we assessed whether any significant psoriasis GWAS signals (*p*-value < 5E-08) mapped to the identified independent NMR GWAS loci. WGS genotype data from 95,372 UK Biobank participants, used in the NMR GWAS, served as the reference panel to calculate linkage disequilibrium (LD) 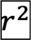 with the index SNP for each overlapping locus.

### Eight psoriasis-related prognostic outcomes

To investigate the associations of lifestyle factors and related metabolomic profiles with the risk of multiple clinical prognostic outcomes among psoriasis patients, we included all-cause mortality, psoriatic arthritis, and six categories of comorbidities[4, 5, 54, 72, 73]: cardiovascular diseases (CVD), obesity-related conditions, gastrointestinal diseases, multiple cancers, mood disorders, and chronic kidney disease. The specific outcomes included in each category are as follows:

(1). Cardiovascular Diseases (CVD): Angina pectoris (ICD-10: I20), acute myocardial infarction (ICD-10: I21), subsequent myocardial infarction (ICD-10: I22), certain current complications following acute myocardial infarction (ICD-10: I23), other acute ischemic heart diseases (ICD-10: I24), chronic ischemic heart disease (ICD-10: I25), hypertension (ICD-10: I10), heart failure (ICD-10: I50), atrial fibrillation (ICD-10: I48), pulmonary embolism (ICD-10: I26), and aortic valve stenosis (ICD-10: I35.0 and I35.2).
(2). Obesity-Related Conditions: Diabetes (ICD-10: E10-E14) and dyslipidemia (ICD-10: E78).
(3). Gastrointestinal Diseases: Crohn’s disease (ICD-10: K50), ulcerative colitis (ICD-10: K51), and nonalcoholic fatty liver disease (ICD-10: K76.0).
(4). Multiple Cancers: Oropharyngeal cancer (ICD-10: C00-C14), esophageal cancer (ICD-10: C15), breast cancer (ICD-10: C50), liver cancer (ICD-10: C22), lung cancer (ICD-10: C33-C34), pancreatic cancer (ICD-10: C25), kidney cancer (ICD-10: C64-C65), bladder cancer (ICD-10: C67), melanoma (ICD-10: C43), non-melanoma skin cancer (ICD-10: C44), mesothelioma (ICD-10: C45), and non-Hodgkin lymphoma (ICD-10: C82-C85).
(5). Mood Disorders: Depression (ICD-10: F32-F33) and anxiety (ICD-10: F40-F48).
(6). Chronic Kidney Disease: Chronic kidney disease (ICD-10: N18).

Psoriatic arthritis diagnoses (ICD-10: L40.5) were obtained from hospital inpatient records (UKB field ID: 41270), and all-cause mortality data were retrieved from national death registries (UKB field IDs: 40000 and 40001).

Six categories of comorbidities were selected for analysis based on prior publications and clinical relevance. We evaluated the associations between receiving a psoriasis diagnosis and the risk of further developing these comorbidities. For each comorbidity, participants with a prevalent diagnosis before the baseline date were excluded. To minimize reverse causality, incident cases occuring within 6 months after the baseline date were also excluded. In each incident comorbidity cohort, participants were categorized based on their psoriasis status as “prevalent” for those diagnosed before the baseline date or during the recruitment period (March 13, 2006 to October 1, 2010) and before the comorbidity diagnosis, or as “control” for those without any recorded psoriasis diagnosis.

Follow-up time for each participant was calculated as the duration from the baseline date to the earliest of the following: comorbidity diagnosis date, death date, or end of follow-up (December 30, 2022). Cox proportional hazards models were used to estimate the association between psoriasis diagnosis and the risk of developing each comorbidity, adjusting for age, sex, BMI, ethnic background, living country, and index of multiple deprivation (IMD). Hazard ratios (HRs) with 95% confidence intervals were calculated at 1-, 2-, 3-, 4-, 5-, 10-, and 15-year follow-up intervals.

The analysis revealed that a prevalent psoriasis diagnosis was a significant risk factor for developing all six comorbidities. Consequently, these comorbidities were included in downstream analyses.

### Hazard ratios for eight prognostic outcomes in patients with psoriasis

The associations of the lifestyle score, LS.mets, and three independent NMR mediators (GlycA, PUFA/MUFA, and creatinine) with the risk of eight clinical prognostic outcomes (all-cause mortality, psoriatic arthritis, and six comorbidities) were evaluated among psoriasis patients (*N* = 15,978). For this psoriasis cohort, the baseline date was defined as the later of the disease diagnosis date or the recruitment date. For each predicted outcome, prevalent cases or those developed the comorbidity/mortality within the 6 months after the baseline were excluded to minimize reverse causality. Follow-up time was calculated from the baseline date to the earliest of comorbidity/mortality diagnosis, death, or end-of-follow-up (October 31, 2022, for psoriatic arthritis and comorbidity diagnosis; November 30, 2022, for all-cause mortality). Cox proportional hazards models were used to assess associations between predictors (the lifestyle score, LS.mets, and three biomarkers) and risk of eight outcomes. Age, sex, BMI, ethnic background, living country, IMD, cholesterol-lowering mediation, and psoriasis PRS were adjusted. For non-CVD outcomes, additional cardiovascular health covariates (stroke, heart failure, and myocardial infarction status) were further adjusted. HRs with 95% confidence intervals were estimated for each predictor-outcome combination.

### Precision-Recall curves

To evaluate the clinical utility of the lifestyle score, LS.mets, GlycA, and PUFA/MUFA in predicting multiple prognostic outcomes, we performed precision-recall (PR) curve analysis among psoriasis patients (*N* = 15,978). PR curves are particularly suited for assessing biomarker performance in imbalanced clinical datasets and have been shown to outperform receiver operating characteristic (ROC) curves in such scenarios[74]. For each outcome, PR curves were generated using event status (1 for incident occurrence and 0 for censoring) as the endpoint and the four markers as predictors. The area under the PR curve (AUPRC) was calculated to quantify predictive performance. Event prevalence (positive proportion) was annotated on each curve to contextualize the results.

### Survival analysis based on GlycA cutoffs

GlycA exhibited the best performance against others in PR curve analyses across seven of the eight outcomes, except for cancers. To further explore its clinical relevance, we stratified psoriasis patients into “high-risk” and “low-risk” groups based on the GlycA cutoff for each outcome and compared disease incidence and survival between two groups. For each clinical prognostic outcome among psoriasis patients, prevalent cases and those occurring within the 6 months after the baseline were excluded. Follow-up time was calculated from the baseline date to the earliest of the event date, death date, or end-of-follow-up (October 31, 2022, for psoriatic arthritis and comorbidity diagnosis; November 30, 2022, for all-cause mortality). For each outcome, the optimal GlycA cutoff was determined as the value that maximized the log-rank test statistics. Patients were stratified into “high-risk” (GlycA above the cutoff) and “low-risk” (GlycA below the cutoff) groups. The log-rank test was then used to compare survival/incidence distributions between these two groups.

### Sensitivity analysis

To evaluate the robustness of our findings, we conducted a series of sensitivity analysis. First, we performed stratified analyses in the incident psoriasis cohort to examine whether the associations between lifestyle, metabolomic profiles, and incident psoriasis risk varied across different subgroups, thereby assessing potential effect heterogeneity.

(1) Age-stratified analysis: participants were grouped into three age categories as 40-50 years (*N* = 91,374), 50-60 years (*N* = 124,578), and 60-70 years (*N* = 153,050).
(2) Sex-stratified analysis: participants were grouped by sex as male (*N* = 176,877) and female (*N* = 193,800).
(3) Genetic risk-stratified analysis: participants were grouped into three genetic risk categories based on the tertiles of the psoriasis PRS as “high” (*N* = 120,583), “middle” (*N* = 120,581), and “low” (*N* = 120,583).

For each subgroup, four multivariate Cox proportional hazards models with varying combinations of covariates were fitted, as described earlier, to assess whether the observed associations remained consistent across strata.

Next, given that UK Biobank participants were middle-aged at recruitment, individuals with persistently unhealthy lifestyles may have developed psoriasis before recruitment. Restricting the analysis to post-recruitment incident cases could underestimate the contributions of lifestyle and related metabolomic profiles to psoriasis risk[29]. To address this, we reassessed the odds ratios of lifestyle status (favorable, intermediate, and unfavorable), lifestyle score, and LS.mets in the prevalent psoriasis cohort, which included cases diagnosed before baseline (prevalent cases, N = 11,053; psoriasis-free controls, N = 486,151). Four multivariate logistic regression models were fitted with psoriasis status as the outcome and four sets of confounders as covariates. Consistent findings in both the incident and prevalent cohorts underscored the importance of upholding to a healthy lifestyle and metabolomic profile in reducing psoriasis risk.

## Data Availability

All data produced are available online at https://doi.org/10.6084/m9.figshare.28747259.v1.

## Acknowledgments

This work was supported by the National Natural Science Foundation of China (Grant Nos. 82272849 to GD), Natural Science Fund for Outstanding Youths in Hunan Province (2023JJ20093 to GD,), and Huxiang Youth Talent Program (Grant Nos. 2023RC3072 to GD, 2024RC3043 to FZ).

