## Supplementary Figures for "The Metabolomic Profile of a Healthy Lifestyle Mediates Psoriasis Risk and Predicts Multiple Comorbidities"

### Supplementary Files

Shiyu Zhang, Yu Meng, Yuming Sun, Yao Yu, Zehao Luo, Daishi Li, Ziyu Guo, Jinchen Li, Furong Zeng, Guangtong Deng, Xiang Chen

Department of Dermatology, Xiangya Hospital, Central South University, Changsha, Hunan, China.

### Supplementary Notes

### Supplementary Figure 1-5

**Supplementary note 1. Detailed definitions and assessments of ten healthy lifestyle components**

1. Smoking status

- UK Biobank field ID: 20116 (smoking status)
- UK Biobank questionnaire/definitions: “Do you smoke tobacco now?” and “In the past, how often have you smoked tobacco?”
- Healthy (1 point): past or never smoker
- Unhealthy (0 point): current smoker

2. Alcohol consumption

- UK Biobank field ID: 1558 (Alcohol intake frequency)
- UK Biobank questionnaire/definitions: “About how often do you drink alcohol?”
- Healthy (1 point): ≤ 4 times/week
- Unhealthy (0 point): Daily or almost daily

3. Physical activity

- UK Biobank field ID: 22036 (At or above moderate/vigorous/walking recommendation)
- UK Biobank questionnaire/definitions: “Indicates whether a person met the 2017 UK Physical activity guidelines of 150 minutes of walking or moderate activity per week or 75 minutes of vigorous activity.”
- Healthy (1 point): ≥ 150 min/week moderate or ≥ 75 min/week vigorous physical activity
- Unhealthy (0 point): < 75 min/week vigorous physical activity

4. Sedentary time

- UK Biobank field ID: 1070 (Time spent watching television)
- UK Biobank questionnaire/definitions: “In a typical day, how many hours do you spend watching TV? (Put 0 if you do not spend any time doing it)”
- Healthy (1 point): < 4 h/day
- Unhealthy (0 point): ≥ 4 h/day

5. Sleep duration

- UK Biobank field ID: 1160 (Sleep duration)
- UK Biobank questionnaire/definitions: “About how many hours sleep do you get in every 24 hours? (please include naps)”
- Healthy (1 point): 7-9 h/day
- Unhealthy (0 point): <7 or >9h/day

6. Fruit and vegetable intake

- UK Biobank field IDs:
  (a) 1319 (Dried fruit intake)
  (b) 1309 (Fresh fruit intake)
  (c) 1289 (Cooked vegetable intake)
  (d) 1299 (Salad/raw vegetable intake)
- UK Biobank questionnaire/definitions:
  (a) “About how many pieces of dried fruit would you eat per day? (Count one prune, one dried apricot, 10 raisins as one piece; put '0' if you do not eat any)”
  (b) "About how many pieces of fresh fruit would you eat per day? (Count one apple, one banana, 10 grapes etc as one piece; put '0' if you do not eat any)"
  (c) "On average how many heaped tablespoons of cooked vegetables would you eat per day? (Do not include potatoes; put '0' if you do not eat any)"
  (d) "On average how many heaped tablespoons of salad or raw vegetables would you eat per day? (Include lettuce, tomato in sandwiches; put '0' if you do not eat any)"
- Healthy (1 point): ≥ 400 g/day (with above four items combined and converted to g/day; 1 portion or tablespoon = 80g)
- Unhealthy (0 point): < 400g/day (with above four items combined and converted to g/day; 1 portion or tablespoon = 80g)

7. Oily fish intake

- UK Biobank field ID: 1329 (Oily fish intake)
- UK Biobank questionnaire/definitions: "How often do you eat oily fish? (e.g. sardines, salmon, mackerel, herring)"
  - *Oily fish include:*
    *Salmon Anchovies*
    *Trout Swordfish*
    *Mackerel Bloater*
    *Herring Cacha*
    *Sardines Carp*

*Pilchards Hilsa*
*Kipper Jack fish*
*Eel Katla*
*Whitebait Orange roughy*
*Tuna (fresh only) Pangas*
*Sprats*

- Healthy (1 point): more than once a week
- Unhealthy (0 point): less than once a week or never

8. Red meat intake

- UK Biobank field IDs:
  (a) 1369 (Beef intake)
  (b) 1379 (Lamb/mutton intake)
  (c) 1389 (Pork intake)
- UK Biobank questionnaire/definitions:
  (a) "How often do you eat beef? (Do not count processed meats)"
  (b) "How often do you eat lamb/mutton? (Do not count processed meats)"
  (c) "How often do you eat pork? (Do not count processed meats such as bacon or ham)"
- Healthy (1 point): ≤ 3 portion/week (with above three items combined and summed up)
- Unhealthy (0 point): > 3 portion/week (with above three items combined and summed up)

9. Processed meat intake

- UK Biobank field ID: 1349 (Processed meat intake)
- UK Biobank questionnaire/definitions: "How often do you eat processed meats (such as bacon, ham, sausages, meat pies, kebabs, burgers, chicken nuggets)?"
- Healthy (1 point): ≤ 1 portion/week
- Unhealthy (0 point): > 1 portion/week

10. Social activity

- UK Biobank field ID: 1031 (Frequency of friend/family visits)
- UK Biobank questionnaire/definitions: "How often do you visit friends or family or have them visit you?" (*Include meeting with friends or family in environments outside of the home such as in the park, at a sports field, at a restaurant or pub*)

- Healthy (1 point): about or more than once a month
- Unhealthy (0 point): less than once a month

### Supplementary Figures

#### Supplementary Figure 1 | Correlation heatmap of 112 lifestyle-associated metabolomic features.

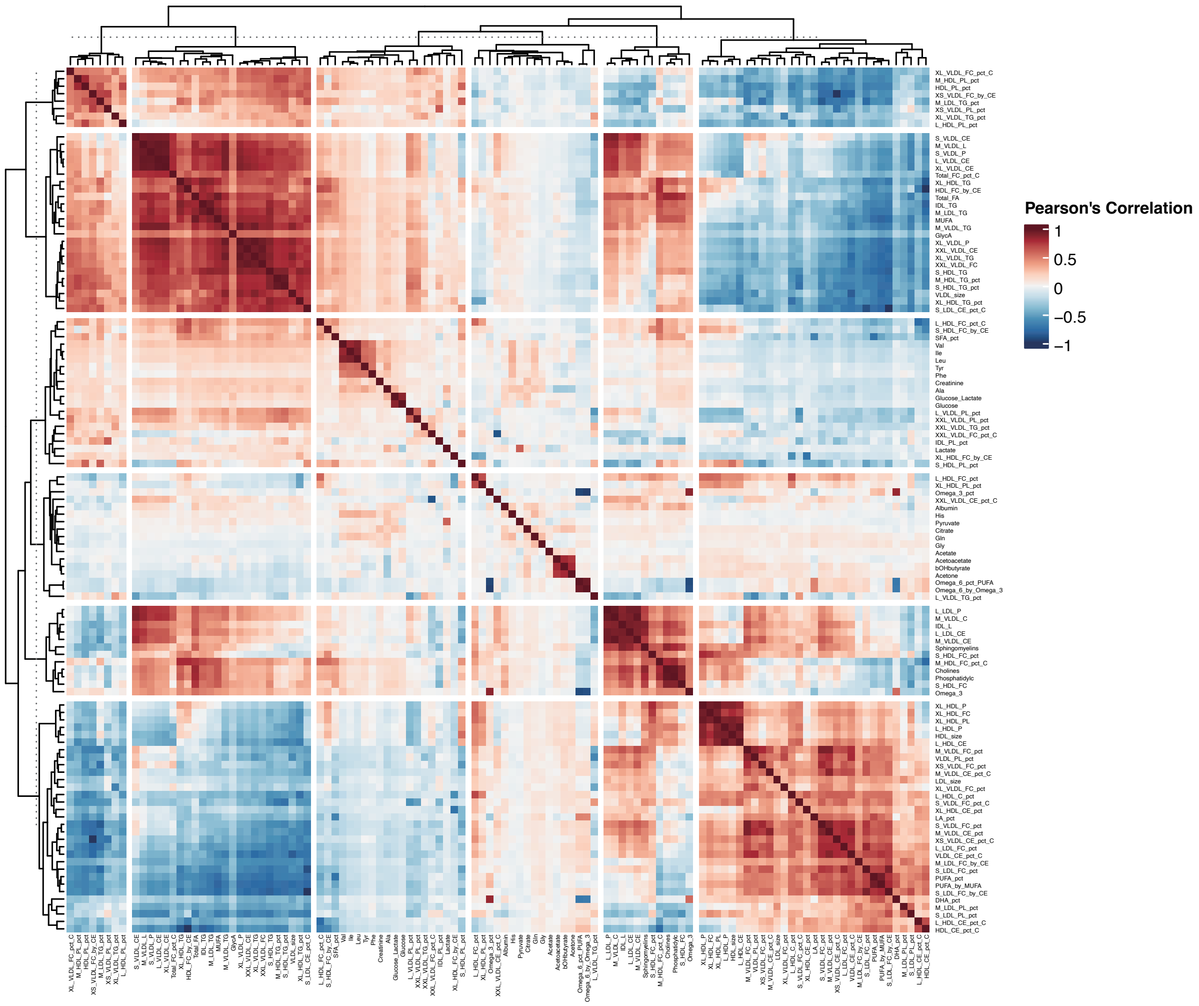


Pearson’s correlation coefficients among the 112 NMR features (N=201,848) were visualized as a heatmap, using a blue-to-red gradient to indicate the direction and magnitude of correlation. The features were grouped into six k-means clusters, determined by the effective number of NMR features (6.037), as described in the Methods.

**Supplementary Figure 2 | Associations of the lifestyle score (LS) and the lifestyle-related metabolomic score (LS.mets) with prevalent psoriasis.**

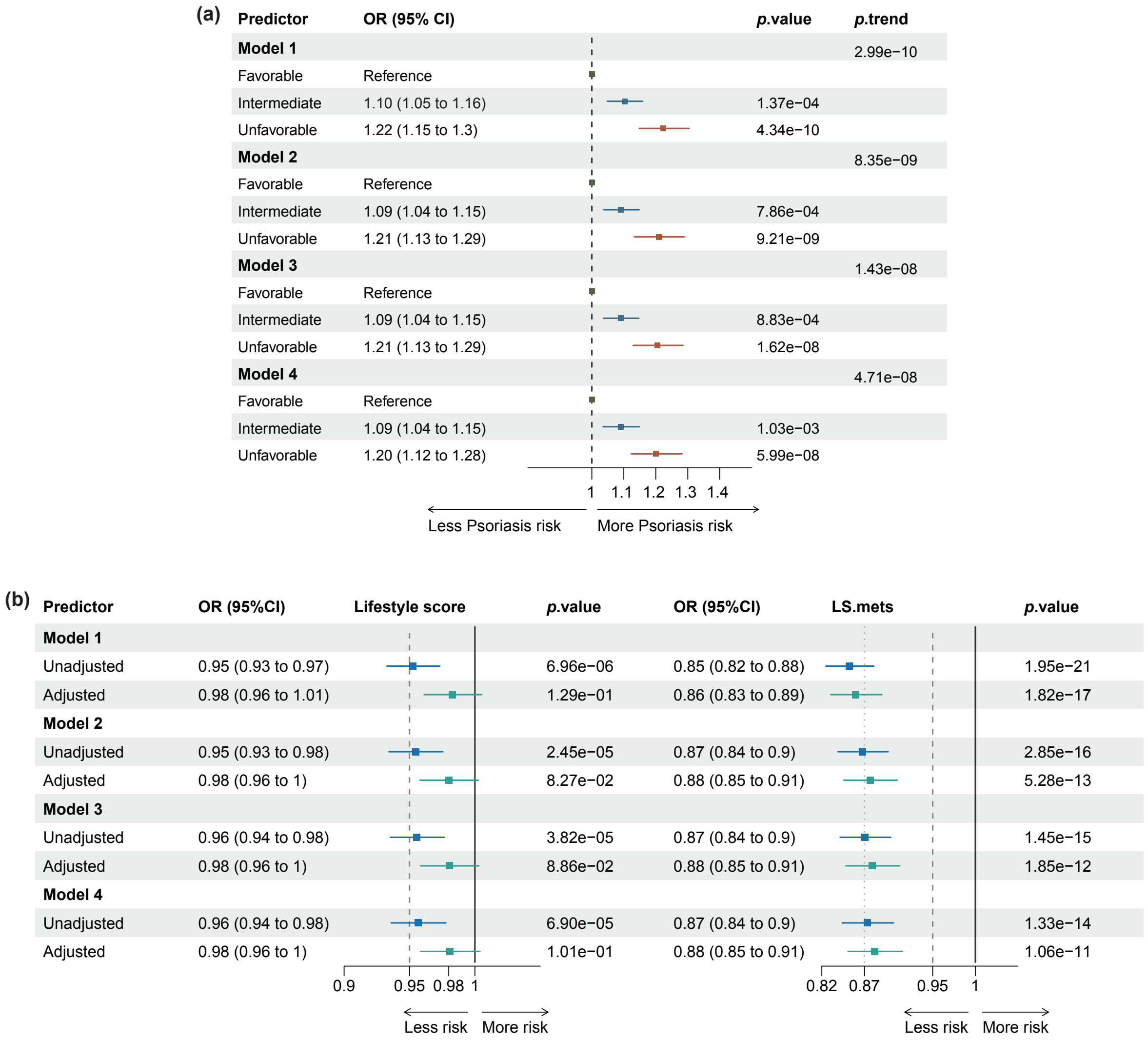


(a) Forest plot displaying odds ratios (ORs) and 95% confidence intervals (CIs) for psoriasis across three lifestyle groups (*N*=375,798), using the favorable group as the reference. Model 1 adjusts for age, sex, and body mass index (BMI). Model 2 additionally adjusts for ethnic background, living country, and index of multiple deprivation (IMD). Model 3 further adjusts for myocardial infarction, stroke, heart failure diagnosis, and cholesterol-lowering medication. Model 4 further adjusts for polygenic risk score (PRS) of psoriasis. The *p*-value for trend was calculated by treating the lifestyle group (ordered as favorable, intermediate, and unfavorable) as a continuous variable.

(b) Odds ratios for prevalent psoriasis diagnosis associated with LS (per point) and LS.mets (per standard deviation) (*N*=187,155). In unadjusted models, each score was evaluated as the individual predictor. In adjusted models, both LS and LS.mets were jointly fitted, thereby isolating their independent associations with prevalent psoriasis.

**Supplementary Figure 3 | Shared genetic architectures of GlycA and PUFA/MUFA with psoriasis in other genomic regions.**

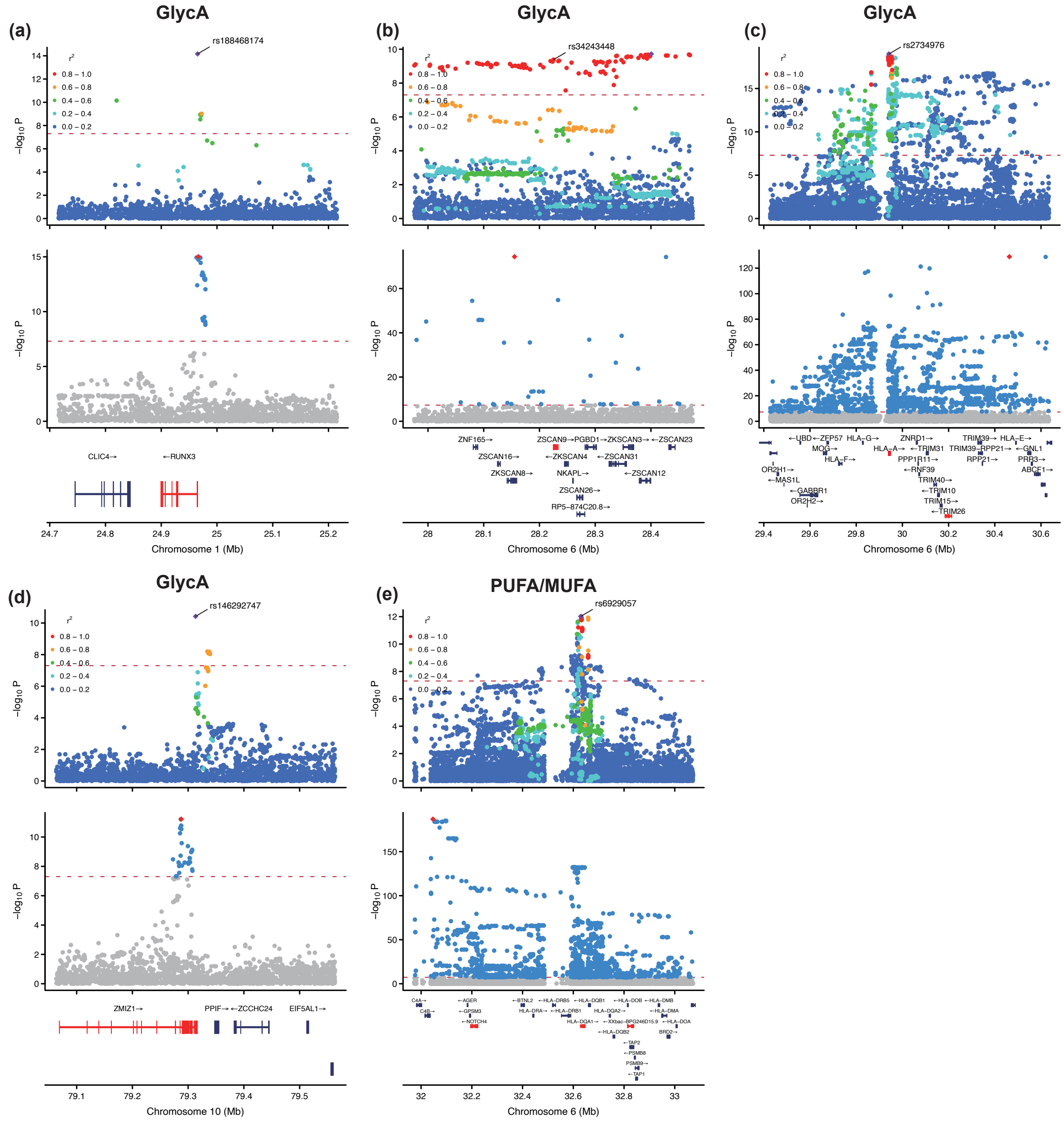


(a) LocusZoom plots showing overlapping GWAS signals of GlycA (top panel) and psoriasis (bottom panel) in the locus: chr1:24,715,206-25,215,206 (hg38 build).

(b) LocusZoom plots showing overlapping GWAS signals of GlycA (top panel) and psoriasis (bottom panel) in the locus: chr6:27,975,324-28,475,324 (hg38 build).

(c) LocusZoom plots showing overlapping GWAS signals of GlycA(top panel) and psoriasis (bottom panel) in the locus: chr6:29,427,768-30,634,245 (hg38 build).

(d) LocusZoom plots showing overlapping GWAS signals of GlycA(top panel) and psoriasis (bottom panel) in the locus: chr10:79,063,270-79,563,270 (hg38 build).

(e) LocusZoom plots showing overlapping GWAS signals of PUFA/MUFA(top panel) and psoriasis (bottom panel) in the locus: chr6:31,972,843-33,071,623 (hg38 build).

Sentinel variants from the NMR biomarker GWAS are labeled, and the nearest genes for independent SNPs (after clumping the NMR GWAS summary statistics) are highlighted in red. The lead SNP in the psoriasis GWAS is also highlighted in red. The red dashed line denotes the genome-wide significance threshold (*p*-value of 5E-08). Linkage disequilibrium (LD) $r^2$ values with the sentinel variant were calculated using whole-genome sequencing data from 95,372 participants (see Methods).

**Supplementary Figure 4 I Prevalent psoriasis diagnosis as an independent risk factor for six comorbidities.**

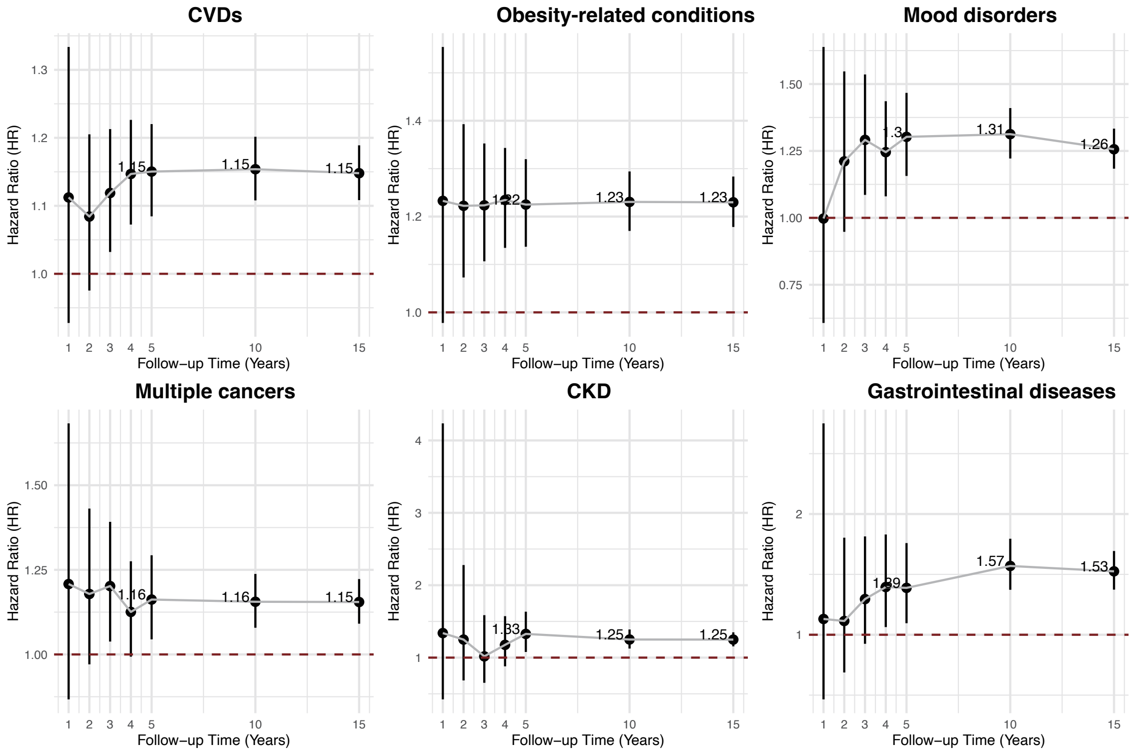


Forest plots illustrating hazard ratios (HRs) for cardiovascular diseases (CVDs) (*N*=437,999), obesity-related conditions (*N*=469,153), mood disorders (*N*=489,403), multiple cancers (*N*=479,769), chronic kidney disease (CKD) (*N*=496,094), and gastrointestinal diseases (*N*=492,818) associated with a prevalent psoriasis diagnosis at different follow-up durations (1 year, 2 years, 3 years, 4 years, 5 years, 10 years, and 15 years). Black circles represent HR estimates with 95% confidence intervals, and the dashed red line (HR=1) indicates the null effect.

### Supplementary Figure 5 | Precision–recall curves for predicting eight clinical outcomes.

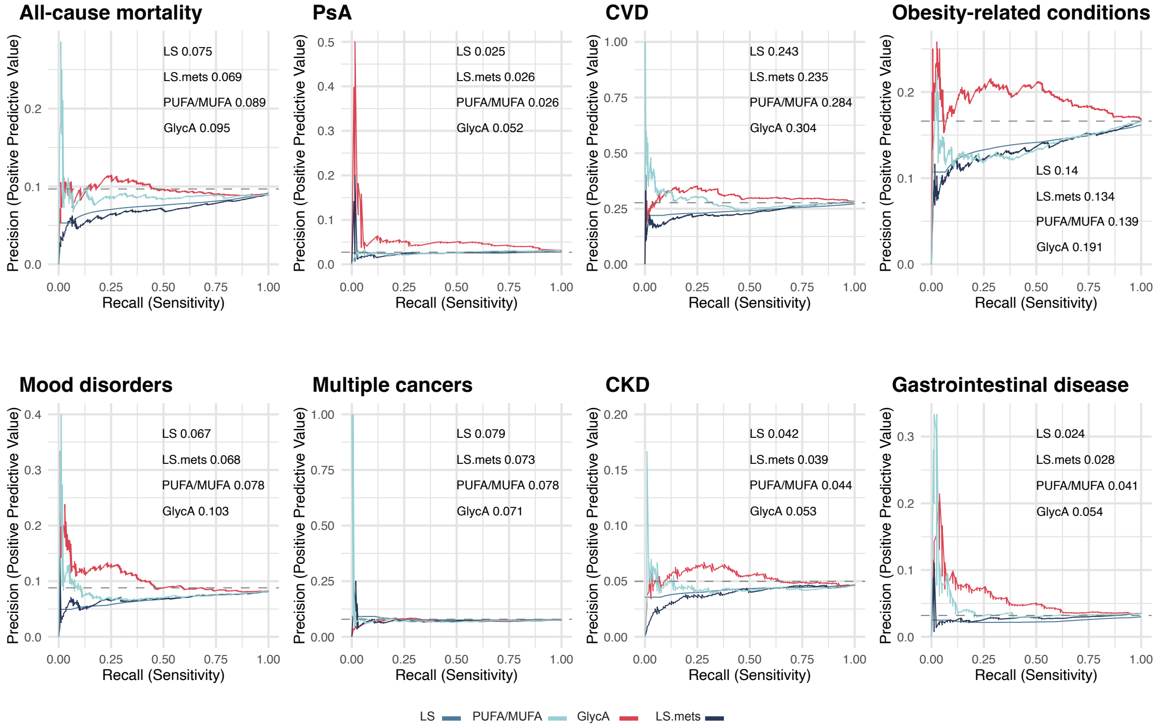


Precision–recall curves compare the predictive performance of the lifestyle score (LS), the lifestyle-associated metabolomic score (LS.mets), PUFA/MUFA, and GlycA across eight prognostic outcomes among psoriasis patients: all-cause mortality (*N*=15,848), psoriatic arthritis (PsA) (*N*=15,091), cardiovascular disease (CVD) (*N*=12,257), obesity-related conditions (*N*=13,940), mood disorders (*N*=15,183), multiple cancers (*N*=15,075), chronic kidney disease (CKD) (*N*=15,718), and gastrointestinal disease (*N*=15,603). The x-axis shows recall (the proportion of true positives correctly identified), and the y-axis shows precision (the proportion of predicted positives that are truly positive).
